# Diverse Relationships Between Antibiotic Resistance and Host Age: A Meta-Analysis Across Antibiotic Classes and Bacterial Genera

**DOI:** 10.1101/2024.02.25.24303263

**Authors:** Lucy E. Binsted, Luke McNally

**Affiliations:** Institute of Ecology and Evolution, School of Biological Sciences, University of Edinburgh, Edinburgh EH9 3FL, United Kingdom

## Abstract

Antimicrobial resistance (AMR) poses an urgent public health challenge. To improve patient outcomes and design interventions we must identify patient characteristics which predict the presence of AMR pathogens. One potential and commonly collected patient characteristic is host age, consensus remains elusive regarding its impact on the probability of infecting pathogens being resistant to antimicrobials. Here, we employ a meta-analysis to consolidate and compare these previous studies and examine the relationship between antibiotic resistance and host age across bacteria and antibiotics. We show that although the probability that infecting bacteria are antimicrobial resistant increases with host age on average, diverse patterns exist across antibiotic classes and bacterial genera, including negative, humped, and U-shaped relationships. We further illustrate, using a compartmental epidemiological model, that this variation is likely driven by differences in antibiotic consumption or incidence of bacterial infection/carriage between age groups, combined with age assortative transmission. These findings imply that empirical antibiotic therapy could be improved by considering age-specific local resistance levels (compared with overall local resistance levels), resulting in improved treatment success and reduced spread of antibiotic resistance. They additionally display consequences of assuming population homogeneity in epidemiological models. Finally, they indicate that the landscape of the already severe resistance crisis is likely to change as the age distribution of the human population shifts.

## Introduction

Antimicrobial resistance (AMR) is a serious and immediate global public health threat ^1^. While the discovery of antibiotics revolutionised modern medicine, rapid emergence and spread of resistance has reduced their efficacy, and with it our capacity to treat and prevent bacterial infections ^2^. This is putting enormous and increasing pressure on healthcare, with an estimated 4.95 million deaths associated with bacterial AMR (antibiotic resistance) in 2019 ^3^. Antibiotic use is the main selective driver for emergence and spread of resistant bacteria ^4^, though where therapeutically justified, is essential for treating infections. As such, in addition to reducing inappropriate antibiotic use, optimisation of antibiotic prescribing is essential.

One way of optimising antibiotic prescribing is to use an antibiotic that the infecting bacteria are sensitive to. In the absence of drug sensitivity results and to prevent the use of broad-spectrum antibiotics which favour the development of resistance ^5^, local resistance levels are often used to predict which antibiotic should be prescribed ^6^. These are detailed in antibiograms, which are generated through monitoring of local resistance probabilities, but do not generally consider patient-specific characteristics. Considering patient characteristics which have an association with the probability that an infecting pathogen is antimicrobial resistant may enable more accurate prediction of the most effective antibiotic ^7^. Many studies have investigated such characteristics, for example age, gender, prior hospitalisations, prior antibiotic treatment, and co-morbidities, but the results are highly variable ^8–12^. This may be because the effect of each individual characteristic is subtle, hence a large sample size would be required to detect it, or that the effect of each characteristic varies across different antibiotics and bacteria. To accurately quantify the effect of a characteristic and variation in this effect, data could be pooled from multiple diverse studies and analysed together in a meta-analysis.

Host age is one of the most commonly investigated risk factors for the proportion of infections caused by resistant (compared with sensitive) bacteria and would be a convenient and simple characteristic for guiding empirical therapy. Moreover, investigating the relationship between resistance probability and host age has important implications for the overall landscape of antibiotic resistance, due to the rapidly increasing age of the human population ^13^. In 2004, an estimated 461 million people were older than 65 but this is set to increase to 2 billion people by 2050 (22% of the population) ^14^. Should older age be associated with an increased ratio of resistant to sensitive infections, an aging population would accelerate the resistance crisis.

There is good reason to suggest resistance probability may vary with host age. Firstly, older people have a less effective immune system ^15^, resulting in more frequent infections. This leads to an increased rate of antibiotic consumption (one of the main drivers for antibiotic resistance ^4^) and more contact with healthcare. Nosocomial (or healthcare-acquired) infections show higher resistance probabilities ^16^ and are often caused by multi-drug resistant pathogens such as MRSA ^17^. Secondly, a weaker immune system could also result in higher bacterial loads, increasing the chance of de novo resistance emergence. Finally, some studies suggest that the frequency of resistance mutations within the host gut microbiome increase with age ^18–20^. Such mutations have the potential of being transferred to pathogenic bacteria via horizontal gene transfer (HGT), thus increasing the likelihood of resistant infections ^21,22^ A recent pre-print, specifically analysing bloodstream infections in Europe for the limited number of antibiotic-pathogen combinations monitored by the ECDC, has also indicated potential associations between AMR probability and age, with varying patterns across antibiotics and bacteria^23^.

The effect of host age may also have consequences for mathematical modelling of antibiotic resistance. Antibiotic resistance is normally modelled using a homogeneous host population ^24^, though with a few notable exceptions ^25–29^. In reality, the human population is highly heterogeneous, a property which affects disease spread and evolution ^30^, and is therefore also likely to affect resistance. Using age-specific parameters based on an understanding of the variation in the probability of antimicrobial resistance with host age is an obvious first step to introduce population heterogeneity within models to make them more refined and realistic.

Through undertaking a meta-analysis of published studies, we firstly aim to investigate the relationship between host age and the probability of an infection being antibiotic resistant and how it varies across different antibiotic classes and bacterial genera. Secondly, we offer explanations for these relationships using age-structured compartmental epidemiological models.

## Results

### Relationship between host age and antibiotic resistance probability

Data for the meta-analysis was gathered through a literature search followed by study selection and data extraction. This process is illustrated in a PRISMA flow diagram (Figure 1) and resulted in 235 studies being included (Supplementary Table 1). From each study, the number of infections in each age category caused by sensitive and resistant bacteria was extracted. Studies testing multiple bacteria or antibiotics were split into different datasets, one for each antibiotic-bacteria combination. Bacteria were grouped by genus (Supplementary Table 3) and antibiotics were grouped by antibiotic class (Supplementary Table 4). The total number of studies, datasets and samples for each antibiotic class and bacterial genus are given in Supplementary Figure 1.

**Figure 1.**
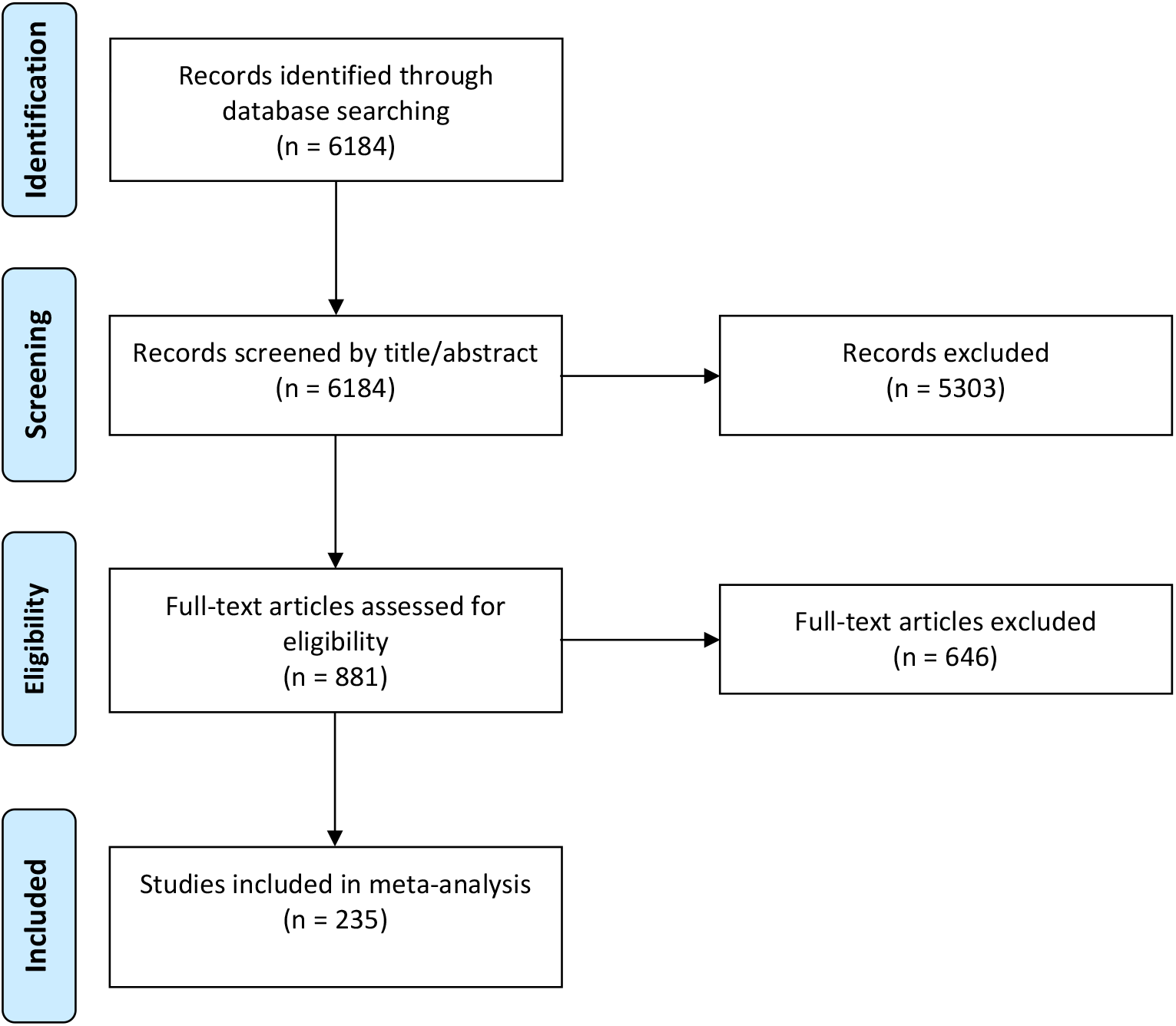
PRISMA flow diagram showing the number of studies included in and excluded from the final meta-analysis. Reasons for exclusion are detailed in Supplementary Table 2.

Firstly, publication bias was assessed by plotting an estimate of the effect size for each study against its standard error, calculated using a binomial regression. This funnel plot was reasonably symmetric, indicating no significant publication bias (Supplementary Figure 2).

Following this, a Bayesian binomial mixed effects model with logit link was fit to investigate the relationship between host age and antibiotic resistance probability. This included age as a fixed effect with a linear (*β*_1_) and quadratic (*β*_2_) term; paper (study) and dataset as random effects on intercept (*β*_0_); and antibiotic class, bacterial genus and class-genus interaction as random effects on intercept (*β*_0_), linear (*β*_1_) and quadratic (*β*_2_) terms.

Although the relationship between resistance probability and age had a saturating relationship (*β*_1 = 1.93 (95% credible interval (95% *C. I*.) = 0.21 − 3.60), *p*_*MCMC*_ = 0.032; *β*_2_ = −1.14 (95% *C. I*. −2.85 − 0.53), *p*_*MCMC*_ = 0.187) on average across class-genus combinations (Figure 2A), there was a large amount of variance in *β*_1_ and *β*_2_ across antibiotic class, bacterial genus and class-genus interaction 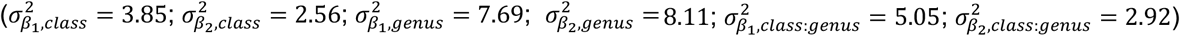 (Figure 2B).

**Figure 2.**
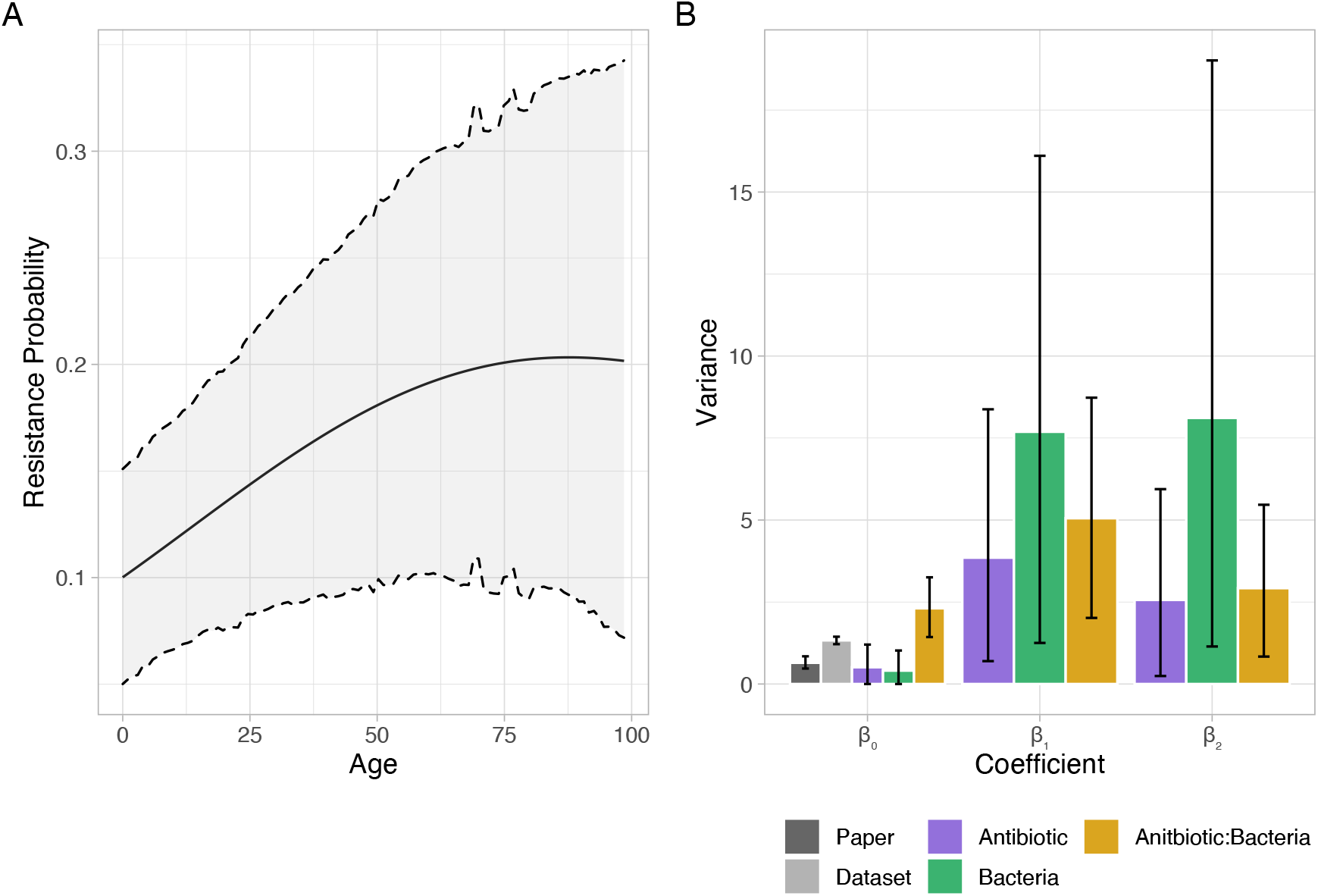
(A) Plot showing the average relationship between antibiotic resistance probability and host age (black line) with 95% credible intervals (grey shading) across all antibiotics and bacteria. (B) Bar graph showing the variance in the relationship between antibiotic resistance probability and age across random effects (paper, dataset, antibiotic, bacteria, and antibiotic-bacteria interaction) for each term (intercept, linear and quadratic).

This variance in *β*_1_ and *β*_2_ across antibiotic class, bacterial genus and class-genus interaction plays out as variation in the shape and magnitude of the relationship between resistance probability and age across both antibiotics and bacteria (Supplementary Figure 3). Variation in shape is summarised in Figure 3A, a plot of the posterior mean for the linear term against the posterior mean for the quadratic term for each antibiotic-bacteria combination. The majority of antibiotic-bacteria combinations had a positive linear and negative quadratic term, resulting in resistance probability increasing then decreasing with age (bottom right quadrant, e.g., Acinetobacter resistance to quinolones). Points above the line *y* = − *x*/2 have a turning point after age 100, so the relationship plateaus at higher ages rather than declining. The next most common relationship had the opposite shape, decreasing then increasing resistance probability with age resulting from a negative linear and positive quadratic term (top left, e.g., Streptococcus resistance to cephalosporins). Furthermore, some combinations had both positive linear and quadratic terms resulting in resistance probability only increasing with age (top right, e.g., Proteus resistance to penicillins), while a couple had both negative linear and quadratic terms resulting in resistance probability only decreasing with age (bottom left, e.g., Pseudomonas resistance to polymyxins). An example of each shape of relationship is shown in Figure 3B, the predicted resistance probability from age 0 to age 100 with 95% credibility intervals. Overall, this shows that the shape of the relationship between antibiotic resistance probability and host age is highly variable across antibiotic classes and bacterial genera.

**Figure 3.**
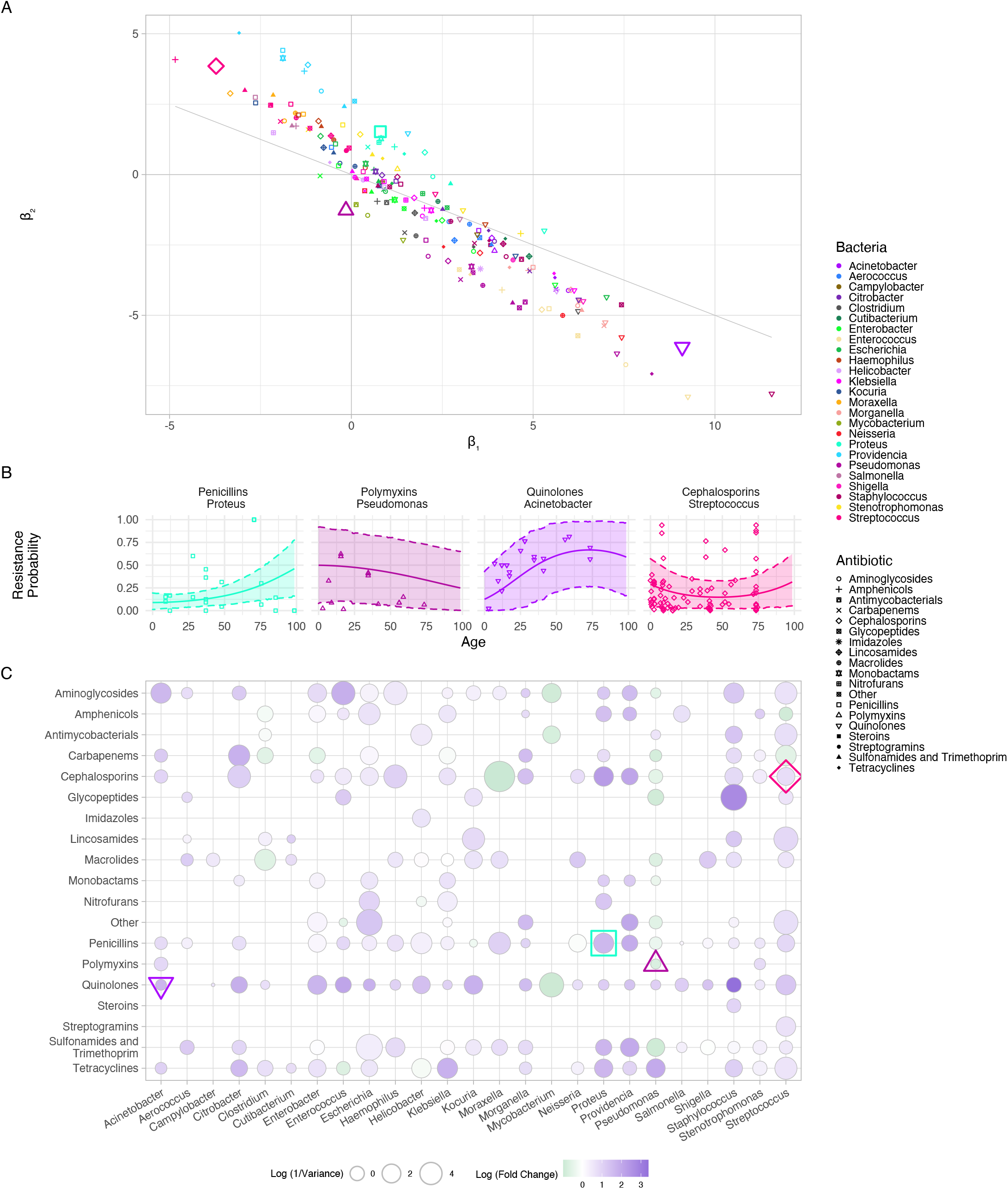
(A) Plot summarising the variation in the shape of the relationship between age and antibiotic resistance probability across antibiotics and bacteria. The posterior mean of the linear term from the MCMCglmm model is plotted against the posterior mean of the quadratic term for each antibiotic-bacteria combination. The grey lines are x = 0, y = 0 and y = –x/2, which separate relationships with different shapes. (B) Plot of the relationship between age and resistance probability for four antibiotic-bacteria combinations, one for each different shape. (C) Plot summarising the variation in the magnitude of the relationship between age and antibiotic resistance probability across antibiotics and bacteria. Colour represents the maximum fold change in resistance probability with increasing age; purple is an increase in resistance probability with age while green is a decrease and white is no change. 1/variance is represented by size. Examples from (B) are highlighted.

Variation in magnitude is highlighted in Figure 3C. This summarises, for each class-genus combination, the predicted resistance probability, and its variance from age 0 to age 100. Firstly, it shows the maximum fold change in resistance probability with increasing age as a measure of how much resistance changes across host age (represented by colour). Secondly the inverse of the sum of the posterior variance in the mean predicted probability of resistance at all ages as a measure of the certainty of the model in the predicted relationship between resistance probability and age (represented by size). Some combinations had a large fold change, for example Staphylococcus resistance to quinolones increased from 0.01 at age 0 to 0.3 at age 74 (fold change = 29.4), while others changed very little, for example Enterobacter resistance to sulfonamides only increased from 0.28 at age 0 to 0.29 at age 45 (fold change = 0.01). In addition, 30 combinations had a fold change below 1, meaning the resistance probability decreased with age. For example, Moraxella resistance to cephalosporins decreased from 0.02 at age 0 to 0.008 at age 55 (fold change = 0.37). The examples from Figure 3B are also highlighted. Overall, this shows that the magnitude of changes in the probability of antimicrobial resistance with host age is also highly variable across both antibiotic classes and bacterial genera.

To investigate the ability of host age to predict the probability of infections caused by resistant versus sensitive bacteria, Receiver Operating Characteristic curves (ROC) analysis was used. This involved fitting individual binomial models of resistance probability on age (including age as fixed effect with a linear and quadratic term) for antibiotic-bacteria combinations with 10 or more age categories. 10-fold cross-validated Area Under the receiver operating characteristics Curve (cvAUC) and 95% confidence intervals was then calculated in each case. An AUC of 0.5 indicates random assignment of resistant and sensitive infections, while >0.7 suggests predictions are good. 59 of the 66 antibiotic-bacteria combinations tested had a cvAUC between 0.5 and 0.7, indicating age was better at predicting resistance probability than random assignment (Supplementary Figure 4). Further, for Staphylococcus resistance to Tetracyclines (cvAUC = 0.790 (0.777 – 0.802)) and Haemophilus resistance to Quinolones (cvAUC = 0.900 (0.861 – 0.938)), age predicted resistance probability well. Interestingly, of the 4 of the 5 antibiotic-bacteria combinations for which age predicted resistance probability worse than random assignment, included Staphylococcus, Haemophilus, Tetracyclines or Quinolones. This, again, highlights the variation in the relationship between resistance probability and host age across antibiotics and bacteria. It is important to note that while the predictive power is generally moderate, these kinds of marginal gains may be important to help slow AMR evolution. Owing to exponential processes in population growth seemingly moderate reductions in the fitness of resistant strains will compound in their effect of time.

**Figure 4.**
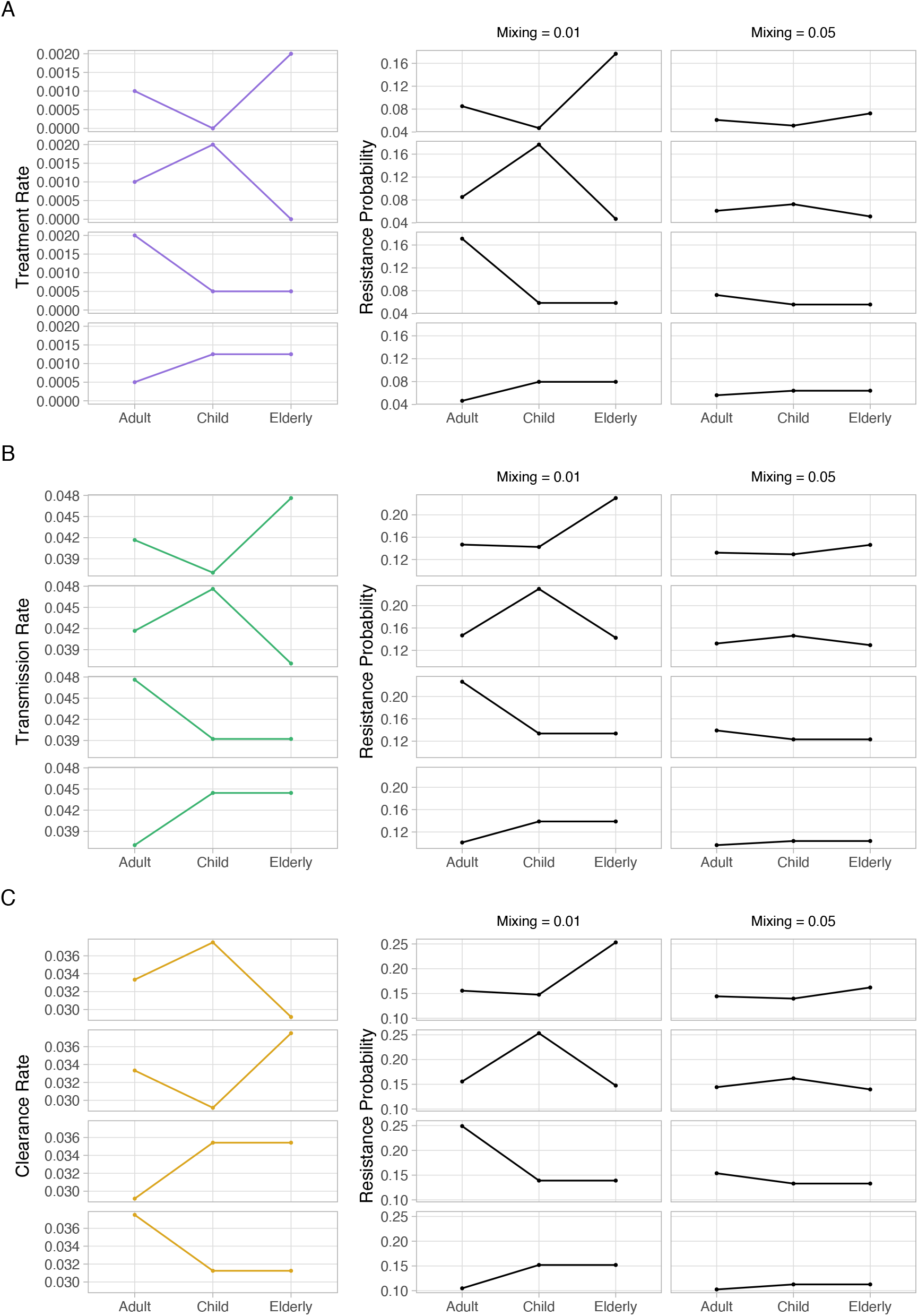
(A) Plots showing how variation in the relationship between age and resistance probability across antibiotics could result from variation in treatment rate between age groups (or sub-populations), and assortative transmission. The first column shows the value assigned to treatment rate in the model, with the second and third columns showing the effect on resistance probability. In the former, between sub-population mixing is lower (0.01), whereas in the latter it is higher (0.05). (B) Plots showing how variation in the relationship between age and resistance probability across bacteria could result from variation in transmission rate (and thus infection/carriage prevalence) between age groups (or sub-populations), and assortative transmission. The first column shows the transmission rate, and the second and third resistance probability with low and high sub-population mixing. (C) Plots showing how variation in the relationship between age and resistance probability across bacteria could result from variation in clearance rate (and thus infection/carriage prevalence) between age groups (or sub-populations), and assortative transmission. The first column shows the transmission rate, and the second and third resistance probability with low and high sub-population mixing. *Multidrug-resistant Acinetobacter baumannii (MDR-AB); Carbapenem-resistant Enterobacterales (CRE); Methicillin-resistant Staphylococcus aureus (MRSA); Vancomycin-resistant Enterococcus (VRE); Extended-spectrum beta-lactamase (ESBL); Multidrug-resistant gram-negative organism (MDR GNO)*.

### Mathematical modelling to explain the relationship between host age and antibiotic resistance probability

Why do we see such high diversity in the age-resistance relationship? With variation occurring both across antibiotics and bacteria, factors driving the relationship must also vary across antibiotics and bacteria. We therefore hypothesised that differences in age-specific treatment rate could underlie variation across antibiotics, while differences in age-specific prevalence (carriage or infection) could underlie variation across bacterial genera. Our justification for this is as follows. Firstly, prescription rates vary across age groups and age-specific prescription rates vary across antibiotics. For example, quinolones are rarely prescribed to children and younger adults, and their usage increases with age ^31^. Since increased antibiotic usage drives the development and spread of resistance by increasing selection pressure ^4^, we would expect resistance to be higher in age groups with high antibiotic usage. Hence, the relationship between resistance probability and age would vary across antibiotics. Secondly, prevalence varies across age groups and age-specific prevalence varies across bacteria. For example, *S. pneumoniae* carriage prevalence is high in children under 5 and decreases with age ^32^. Higher prevalence increases the likelihood of co-infection (where an individual is simultaneously infected with two strains of bacteria: one sensitive and the other resistant), thus increases within-host competition. In the presence of antibiotics, sensitive bacteria are eliminated leading to competitive release (expansion of a species when the competitor for its niche is removed ^33^) of resistant bacteria. Hence, we would expect resistance to be higher in age groups with high prevalence and for this to vary across bacteria. In order to maintain differences in resistance probability between age groups, we proposed there must also be age-assortative transmission, where individuals transmit bacteria at a higher rate to those within their age group. Different age groups therefore represent distinct but interacting sub-populations within a larger heterogeneous overall population ^34^.

To test these hypotheses, a susceptible-infected compartmental model was developed (Supplementary Figure 5). This consisted of an uninfected host class (*U*) and three infected host classes (antibiotic-sensitive infection, *I*^*S*^; antibiotic-resistant infection, *I*^*R*^; and co-infection, *I*^*SR*^) in three age categories. Transmission of either sensitive or resistant bacteria to uninfected individuals occurred at rate *β*, with resistant bacteria incurring a transmission cost of resistance (*ct*) leading to a reduction in transmission rate. Subsequent transmission of resistant bacteria to sensitive-infected individuals, or sensitive bacteria to resistant-infected individuals also occurred at rate *β* with a transmission cost of resistance (*ct*), potentially leading to co-infection. A small proportion (*m*) of contacts were with individuals from the other two age categories in equal amounts, while a larger proportion (*S* − *m*) of contacts were with individuals from the same age category as the focal individual. Thus, most bacteria came from infected individuals in the same age category, with a small proportion coming from infected individuals in the other two age categories. All bacteria were cleared by the immune system at rate *γ*, and sensitive bacteria were additionally cleared by antibiotics at rate *ατ*, where *α* was antibiotic clearance rate and *τ* was sub-population-level antibiotic treatment rate. *β, γ*, and *τ* were specific to each age group.

To determine whether age-specific treatment rate could lead to the observed relationships between resistance probability and age, treatment rate (*τ*) was varied across the three age groups while keeping all other parameters the same. As predicted, high treatment rate in an age group resulted in high resistance probability and low treatment rate in low resistance probability (Figure 4). However, these differences in resistance probability between age groups were only maintained when between age-group transmission was low (mixing = 0.01) and began to level out when between age-group transmission was higher (mixing = 0.05). To determine whether age-specific prevalence could lead to the observed relationships between resistance probability and age, either transmission rate (*β*) or immune clearance rate (*γ*) was varied across the three age groups while keeping all other parameters the same. With this parameter set, high prevalence (as a result of high transmission or low immune clearance) in an age group resulted in high resistance probability, and low prevalence in low resistance probability. Again, the differences in resistance probability between age groups were only maintained when between age-group transmission was low. These results indicate that age-specific treatment rate is likely to at least partly explain variation in the relationship between the probability of antimicrobial resistance and host age across antibiotics, while age-specific prevalence is likely to at least partly explain variation in this relationship across bacterial genera.

Further to these results, the models also display stable coexistence of resistant and sensitive bacteria at the host population level. That is, resistance probability does not tend to either 0 or 1 at equilibrium but settles somewhere in between. Coexistence is observed empirically but is often challenging to capture in simple mathematical models^25,35^. This suggests that including elements of host population variability, such as age, in mathematical models may improve their accuracy and thus their ability to make useful predictions regarding AMR dynamics and the effects of possible interventions.

## Discussion

By fitting a model to age-specific resistance data from 235 different studies, we quantified the relationship between age and resistance probability and found there to be high variation across both bacteria and antibiotics. Of note, there was a particularly strong relationship between age and resistance probability for quinolone antibacterials, a result which has been described previously ^36^. We also found, using ROC-AUC analysis, that age was moderately predictive of resistance probability. Age was able to predict resistance probability well for Staphylococcus resistance to tetracyclines and Haemophilus resistance to quinolones and was better than random for most other combinations assessed. While this may be a moderate effect for individual patients, such effects could help reduce selection and slow resistance evolution on a population-level. We further constructed a mathematical model to show that age-specific treatment rate and age-specific prevalence could underlie the relationship between resistance probability and age, and its variation across antibiotics and bacteria respectively.

Higher treatment rates and increased prevalence in older compared with younger people may also contribute to the overall age-resistance relationship (i.e., increasing resistance probability with age). In general, older people have a less effective immune system ^15^, resulting in more frequent and more severe infections ^37^. This means they are typically prescribed antibiotics at higher rates than healthy adults ^38^. Furthermore, additional health declines also tend to result in increased contact with healthcare settings. The latter has not been discussed but is worth noting since inpatient (particularly hospital-acquired) infections show higher resistance probabilities ^16^ and are often caused by multi-drug resistant pathogens such as MRSA ^17^. Additionally, biased population mixing within a hospital or long-term care facility is likely to exaggerate this effect, as shown in our epidemiological model. Finally, resistance mutations within the host gut microbiome could accumulate throughout life and be acquired by pathogenic bacteria via horizontal gene transfer (HGT) ^39^.

This research argues that using age-specific antibiograms to inform empirical antibiotic therapy and considering different age groups when targeting resistance interventions may be more effective than current strategies. Such a regimen would require a larger amount of data for age-stratified resistance probability in clinically relevant antibiotics and bacteria, though patient age is usually a readily available and easily recorded piece of information. To further improve targeting of empirical therapy and resistance interventions, this idea could be extended to account for other factors which may correlate with the probability of infections being caused by resistant versus sensitive bacteria. For instance, distinguishing between infections acquired in community and healthcare settings may be important, as a higher proportion of infections acquired in hospitals exhibit resistance compared to those acquired in the community ^40,41^.

In addition, the ability of our age-structured mathematical models to capture real-life resistance dynamics suggest that age-structure should be considered in future models, rather than a homogenous host population. Our model not only displays a relationship between resistance probability and age with variation across antibiotics and bacteria, but also stable coexistence of resistant and sensitive bacteria, a property not usually observed in simple mathematical models of antibiotic resistance ^25,35^.

From this research, we conclude that the changing age structure of the human population could significantly alter the makeup of antibiotic resistance. More accurate estimates for carriage, infection and resistance rates by age are urgently required to determine exactly what this will entail for each antibiotic class and bacterial genus.

## Materials and Methods

### Study selection

Articles which investigated antimicrobial resistance across different age groups and indexed before 02/10/2020 were identified through a literature search in the PubMed database using the following search terms: ((age[Title/Abstract]) OR (ages[Title/Abstract]) OR (aged[Title/Abstract]) OR (aging[Title/Abstract]) OR (ageing[Title/Abstract]) OR (old[Title/Abstract]) OR (older[Title/Abstract]) OR (elderly[Title/Abstract]) OR (elder[Title/Abstract]) OR (geriatric[Title/Abstract])) AND (adult[MeSH Terms]) AND (drug resistance, microbial[MeSH Terms]).

Subsequently, these articles were filtered. Review articles, case reports, comments, guidelines, articles not written in English and articles relating to fungal or viral antimicrobial resistance were excluded based on a title screen. Remaining texts were then screened based on abstract, with reasons for exclusion detailed in Supplementary Table 2. Studies which passed the abstract screen were assessed for eligibility through screening of the full text. Studies for which the full text could not be accessed were excluded. Studies were considered eligible if the number of infections caused by antibiotic-sensitive bacteria and the number of infections caused by antibiotic-resistant bacteria was detailed for more than one age category. In addition, the specific bacterial species or genus causing the infection must have been documented, as well as the specific antibiotic or class resistance was tested for. Reasons for exclusion are detailed in Supplementary Table 2 and the number of articles removed at each stage is shown in Figure 1. Papers were excluded at the point in which they met an exclusion criterion, and this was reported as the reason for exclusion. Presence of additional exclusion criteria were not noted. A list of studies included in the final meta-analysis is given in Supplementary Table 1 and details are provided in the supporting dataset.

### Data collection and processing

For every paper, a separate dataset was created for each antibiotic-bacteria combination. Datasets with no resistant infections were excluded and those which varied by other variables (e.g., gender, location, date) were combined. One exception was made ^42^, which had two datasets for methicillin resistance in S. aureus varying by date but using different age ranges. For each dataset, the minimum age (*x*_*min*_), maximum age (*x*_*max*_), number of sensitive infections and number of resistant infections was extracted for each age category. The number of sensitive and resistant infections were either taken directly, calculated from percentage resistance or extracted from graphs using WebPlot Digitizer ^43^. If an age category spanned less than one year (e.g., 0-2 months), it was combined with the next age category. This data is provided in the supporting dataset. The median of an age category (*x*_*median*_) was used as an approximation for all individuals in that age category:

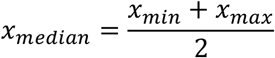

In some cases, the lowest age category did not have a lower bound. Here, the median age of the age category was predicted using a linear regression of median age on maximum age. This regression was done using all age categories that had both an upper and lower bound:

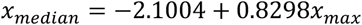

Similarly, there were instances where the highest age category did not have an upper bound. In these cases, the median age was predicted using a linear regression of median age on minimum age. Again, this regression was done using all age categories that had both an upper and lower bound:

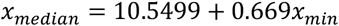

Bacteria were grouped according to genus (Supplementary Table 3), and antibiotics were grouped using the World Health Organization Collaborating Centre for Drug Statistics Methodology ATC/DDD index (Supplementary Table 4). The total number of papers, datasets and samples for each antibiotic-bacteria combination are shown in Supplementary Figure 1.

### Publication bias

To check for publication bias, the effect size of each paper was plotted against 1/Standard Error in a funnel plot (Supplementary Figure 2). Effect size and standard error were estimated for each paper using a binomial regression which included age as a fixed effect with a linear term (*β*: Estimate = effect size, Std.Error = standard error).

### Binomial generalised linear mixed model

To investigate the relationship between resistance probability and host age in the collected data, the R package MCMCglmm ^44^ was used to fit a binomial generalised linear mixed model with logit link. This method uses an MCMC algorithm to determine the most likely parameters given the observed data and prior distribution. A diffuse prior was chosen using an inverse Wishart distribution for the random effects and a normal distribution for the fixed effects. The model included age as a fixed effect with a linear and quadratic term; paper (study) and dataset as random effects on intercept (*β*_0_); and antibiotic class, bacterial genus, and class-genus interaction as random effects on intercept (*β*_0_), linear (*β*_1_) and quadratic (*β*_2_) terms. Antibiotic class, bacterial genus and class-genus interaction were included as random effects rather than fixed effects since some groups had very small sample sizes. In addition, there were many different antibiotic classes (total = 19), bacterial genera (total = 25), and class-genera combinations (total = 191). Geographical location, method of acquisition, study date and infection status could not be included in the model as additional random effects since most datasets could not be classified into a single category, thus would have resulted in a large amount of data loss. The effect of these characteristics on resistance probability could be investigated independently of age but would require further data extraction.

To determine whether inclusion of the quadratic term and random effects was justified, the Deviance Information Criteria (DIC) for the model described above was compared to (a) the same model without quadratic terms and (b) the same model without paper, dataset, class, genus, or class-genus interaction as random effects. The Deviance Information Criterion (DIC) is a Bayesian method for model comparison akin to the Akaike Information Criterion. The DIC was lower when the quadratic term was present (3929477 compared with 3929571), justifying its inclusion. Similarly, the presence of paper, dataset, class, genus, and class-genus interaction as random effects in the quadratic model greatly decreased the DIC (3929477 compared with 3930481), indicating that inclusion of these random effects was also justified. This was additionally supported by histograms showing the distribution of the variance of each random effect (Supplementary Figure 6) and the fact that the lower bound of the credible interval for each random effect was far from zero (Figure 2B). Model convergence was assessed qualitatively by plotting the trace and density of the posterior distribution for fixed (Supplementary Figure 7) and random (Supplementary Figure 8) effects, and quantitatively by estimating a Potential Scale Reduction Factor (PSRF) using Gelman and Rubin’s convergence diagnostic. This involves comparing the MCMC chain of the original model to MCMC chains from two additional quadratic mixed effects models with different starting values. Good convergence in MCMCglmm models is essential for obtaining valid, precise, and reliable parameter estimates, ensuring accurate statistical inferences, and avoiding biases. Traces of the posterior distributions were smooth and random without obvious patterns or trends, while density plots were unimodal, symmetric, and stable. This indicates that the model had good convergence. This was supported by a Potential Scale Reduction Factor (PSRF) estimate and upper confidence interval of 1.

The posterior mean, 95% credibility intervals and variance, for the intercept (*β*_0_), linear (*β*_1_) and quadratic (*β*_2_) terms across all antibiotic classes and bacterial genera and for each antibiotic class bacterial genus combination were calculated from their posterior distributions. These are shown in Figure 2B and Figure 3A.Resistance probability at age 0 to age 100 was calculated for each MCMC iteration using the posterior distribution of the intercept (*β*_0_), linear (*β*_1_) and quadratic (*β*_2_) terms across all antibiotic classes and bacterial genera and for each antibiotic class bacterial genus combination. The mean resistance probability, 95% credibility intervals, and variance for each age were then calculated over all iterations. This is shown in Figure 2A and Supplementary Figure 3). For each antibiotic class bacterial genus combination, the age at which resistance probability was minimum and the age it was maximum was determined and the fold change in resistance probability between these ages was calculated. This is shown in Figure 3C.

### ROC-AUC analysis

To assess the ability of the model to predict antibiotic resistance based on age, we used Receiver Operating Characteristic curves (ROC) analysis with a 10-fold cross-validation approach to avoid overfitting (Supplementary Figure 4). For each class-genus combination with a large enough sample size (> 9 age categories), data was split into 10 subsets (“folds”). The Area Under the ROC Curve (AUC) was then calculated for each subset in turn, using predictions from a model (binomial generalised linear mixed model with logit link including age as a fixed effect with a linear and quadratic term) trained on the other 9 subsets. Folds were defined by splitting the dataset using stratified random sampling, ensuring equal representations of the binary outcomes in each fold. The function ci.cvAUC() from package cvAUC ^45^ was used to retrieved the mean cross-validated AUC and its associated confidence interval.

### Mathematical model

In order to investigate factors which could be influencing the variation in the age-resistance relationship across antibiotics and bacteria, a susceptible-infected compartmental model was developed. This comprised 4 classes of individual (uninfected, *U*; antibiotic-sensitive infected, *I*^*S*^; antibiotic-resistant infected, *I*^*R*^; and co-infected, *I*^*SR*^). Uninfected individuals were infected with either sensitive bacteria at rate *β* (transmission rate without cost of resistance), or resistant bacteria at rate *β*(1 − *ct*) (transmission rate with cost of resistance). We assumed no simultaneous acquisition of sensitive and resistant bacteria. Instead, sensitive infected individuals were infected with resistant bacteria at rate *β*(1 − *ct*) and resistant infected individuals were infected with sensitive bacteria at rate *β*, resulting in coinfection. For each age group, a proportion of transmission (*m*) came equally from infected individuals in the other two age groups, with the rest (1 − *m*) coming from infected individuals in the same age group. Sensitive-infected individuals, returned to the uninfected class at rate *γ* + *ατ*, where *γ* represents clearance of bacteria by the immune system, *α* represents clearance of bacteria by antibiotics and *τ* represents sub-population-level antibiotic treatment rate. Resistant- and co-infected individuals, returned to the uninfected class at rate *γ*, since resistant bacteria are not cleared by antibiotics. Lastly, co-infected individuals became resistant-infected at rate *ατ*, representing antibiotic clearance of sensitive bacteria. *β, γ*, and *τ* were specific to each age group, while *ct* and *α* were the same for all ages. A diagram of the model is displayed in Supplementary Figure 5. This model is simple and does not attempt to accurately describe real-life, but to provide proof of concept. The model assumes that between population mixing occurs equally, rather than using more complicated age-based contact networks to estimate the amount of between age group transmission ^34^.

Baseline parameters for the model were established using community-acquired carriage of *Streptococcus pneumoniae* in the UK and resistance to penicillins (Supplementary Table 5). Immune clearance rate (*γ*) was calculated by 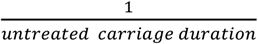, with untreated carriage duration estimated to be 30 days (∼50 days in children, ∼20 days in adults and elderly ^46,47^). Transmission rate (*β*) was calculated by 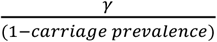, with carriage prevalence estimated to be 0.2 (∼0.4 in Children, ∼0.1 in adults and elderly ^32,48–51^), and transmission cost of resistance (*ct*) was estimated as 0.05 ^52–54^. Antibiotic clearance (*α*) was calculated by 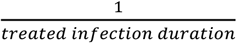, with treated infection duration estimated to be 1 day ^55^. Treatment rate (*τ*) was estimated using data from the European Centre for Disease Prevention and Control (ECDC) on the primary-care consumption of penicillins in the UK in 2019 ^56^. 11.12 Defined Daily Doses (DDDs) per 1000 persons per day was converted to treatment rate by assuming that 10 DDDs comprise 1 treatment course ^57^. Corresponding data from the ECDC Surveillance Atlas was used to approximate penicillin non-susceptibility in *S. pneumoniae*, also in the UK in 2019 ^58^.

The model was written as ordinary differential equations and solved using the deSolve package in R. Starting values for sensitive infections was 90% of the estimated carriage prevalence (0.18), for resistant infections was 10% of the estimated carriage prevalence (0.02), and for co-infections was 0. The model was run for 3650 days (10 years), to ensure it reached a stable equilibrium, and the final resistance probability was taken. The model was first run for a single population, and the equilibrium resistance probability compared with real-life surveillance data.

Next the model was run for three distinct but interacting populations (child, x; adult, y; and elderly, z) while varying single parameters (*τ, γ*, or *β*) between them, to determine the impact on resistance probability. For each of these cases, the model was run with two different values for *m*, which represents the degree of between population mixing. Parameter values, resistance probability, and infection prevalence are given in Supplementary Table 6. Below are the differential equations describing the rate of change of uninfected, single-infected, and co-infected individuals in different sub-populations (child, x; adult, y; elderly, z):

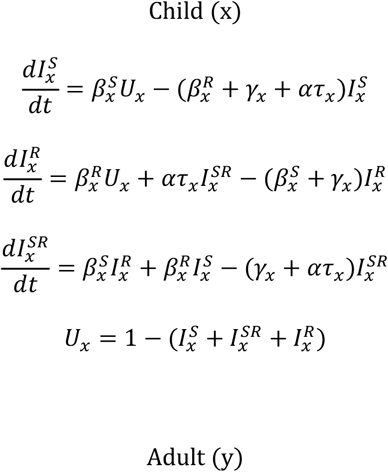

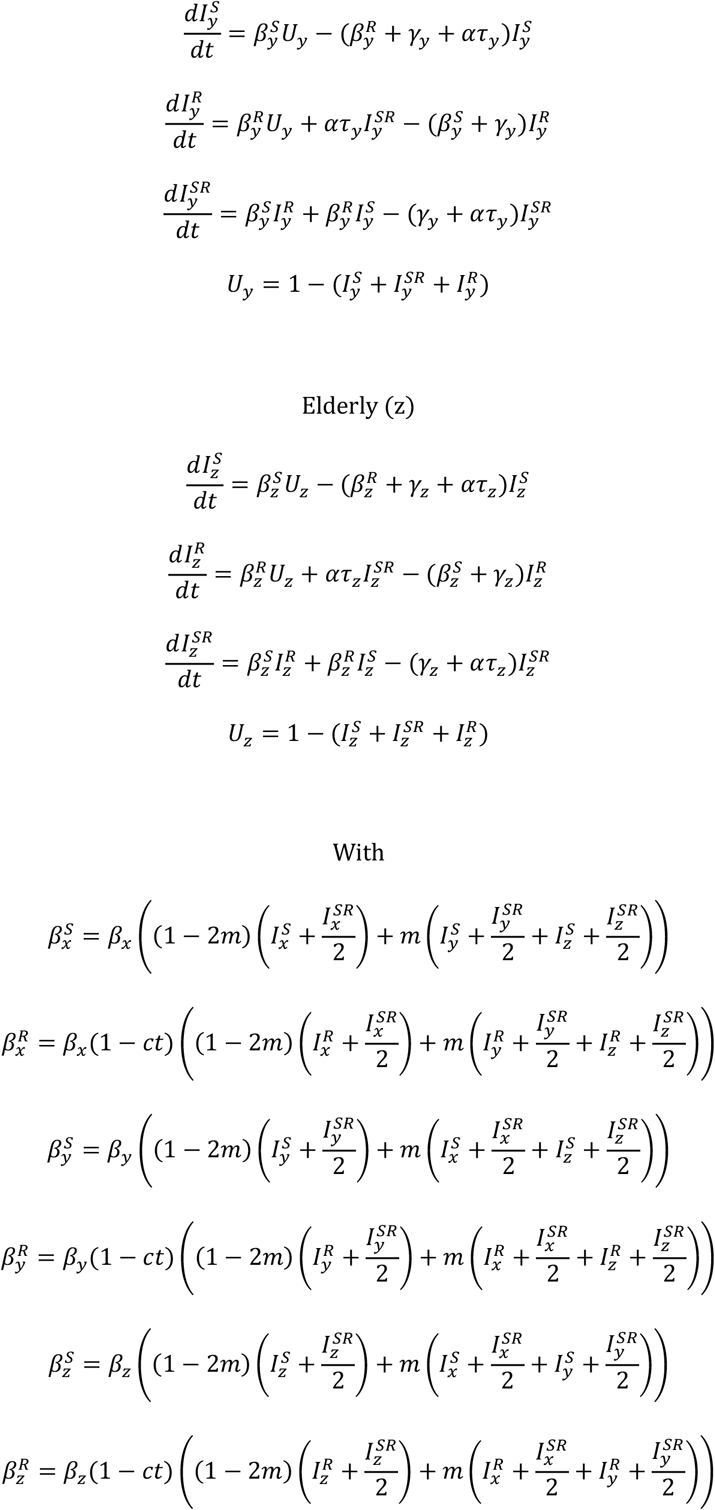

## Data Availability

All data produced in the present work are contained in the manuscript.

https://github.com/LucyEmma22/Age-AMR

