## Supplementary Figures and Tables for "Diverse Relationships Between Antibiotic Resistance and Host Age: A Meta-Analysis Across Antibiotic Classes and Bacterial Genera"

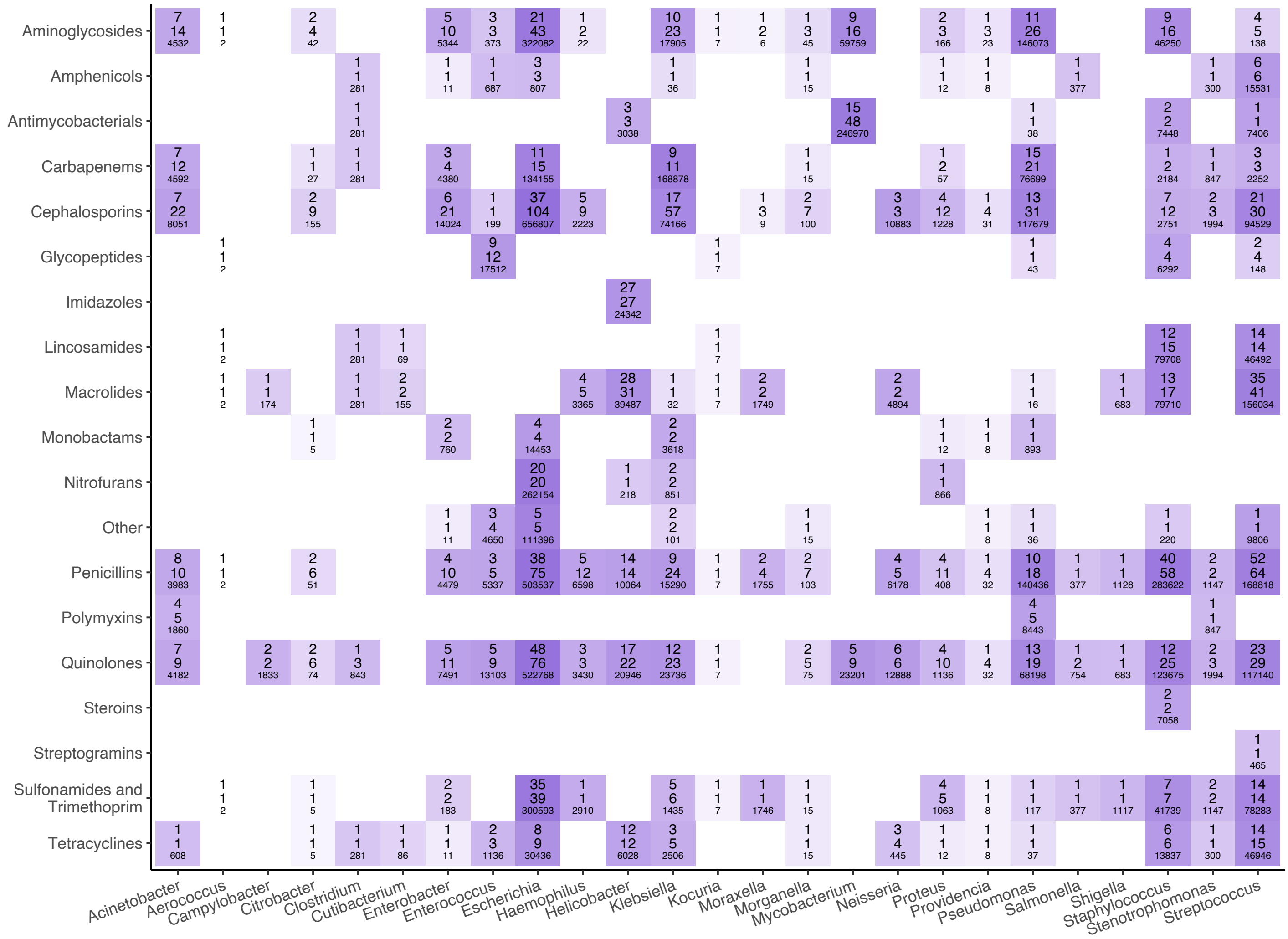


**Fig. S1.** Plot showing the total number of studies (top), datasets (middle) and samples (bottom) for each antibiotic-bacteria combination. Darker squares represent a larger total sample size.

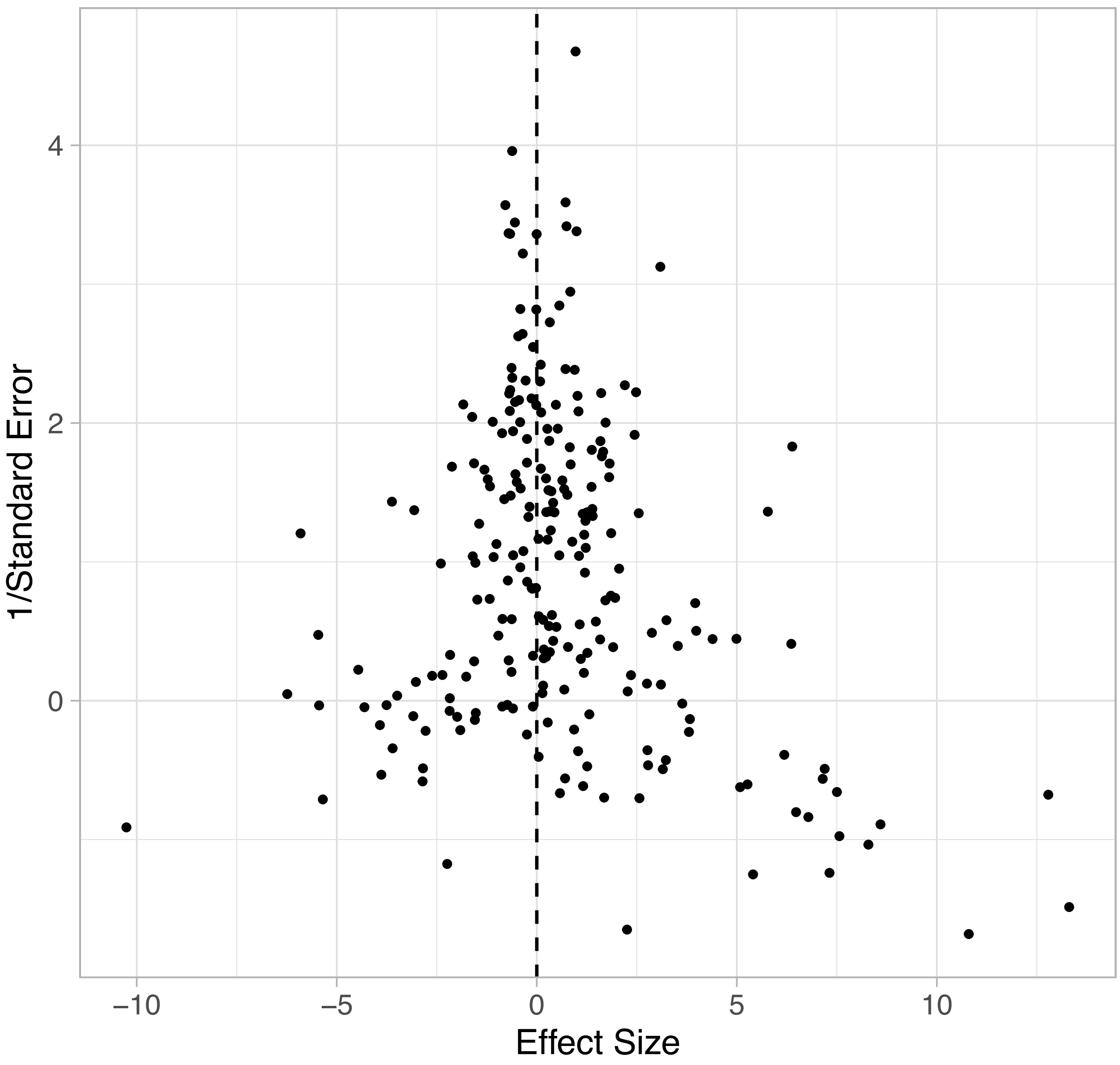


**Fig. S2.** Funnel plot to assess publication bias. Each point represents a paper. Effect size and standard error for each paper was calculated using a simple linear regression of resistance probability on age.

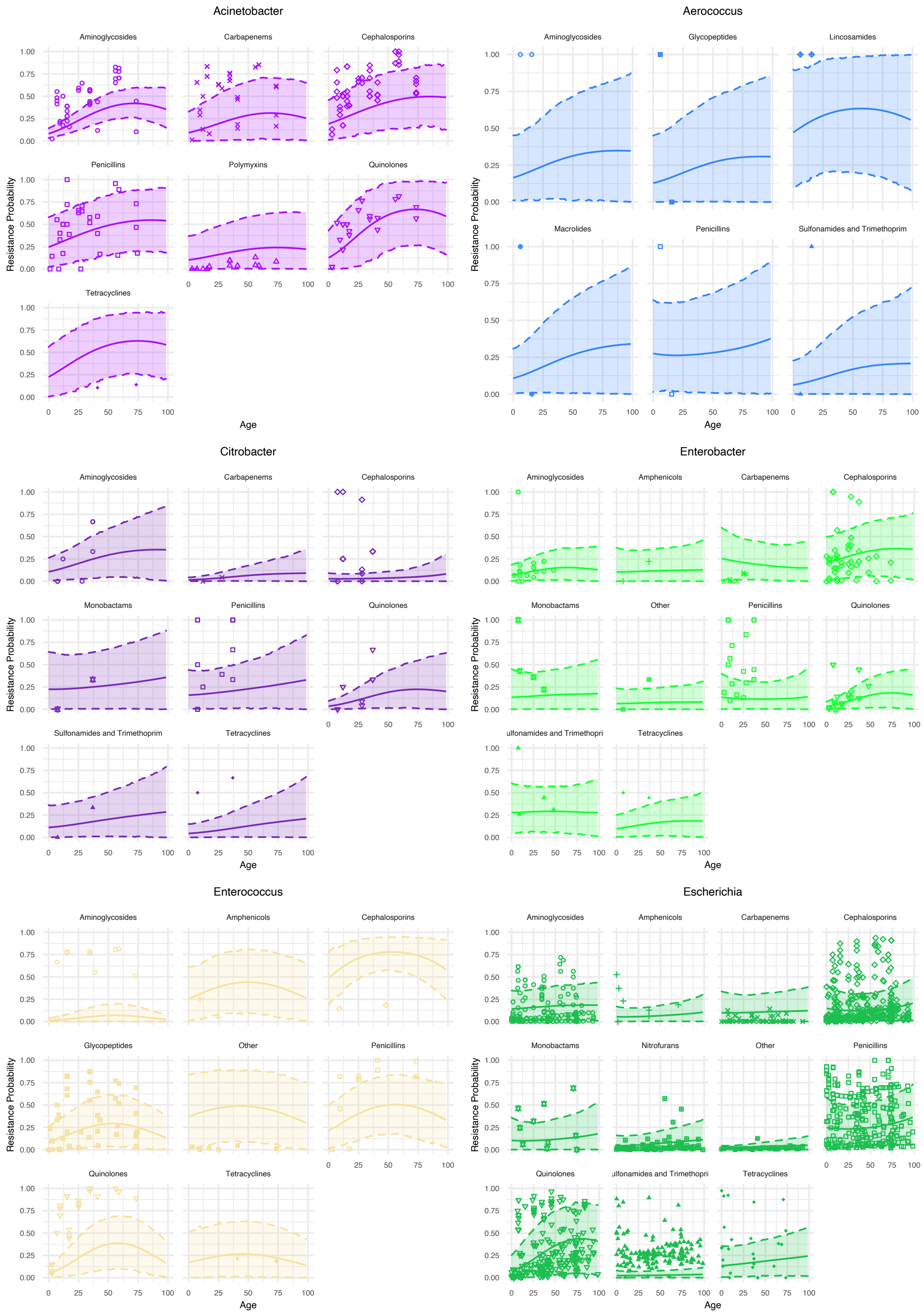

Acinetobacter
Aminoglycosides
Carbapenems
Cephalosporins
Penicillins
Polymyxins
Quinolones
Tetracyclines
Aerococcus
Aminoglycosides
Glycopeptides
Lincosamides
Macrolides
Penicillins
Sulfonamides and Trimethoprim
Citrobacter
Aminoglycosides
Carbapenems
Cephalosporins
Monobactams
Penicillins
Quinolones
Sulfonamides and Trimethoprim
Tetracyclines
Enterobacter
Aminoglycosides
Amphenicols
Carbapenems
Cephalosporins
Monobactams
Other
Penicillins
Quinolones
ulfonamides and Trimethopri
Tetracyclines
Enterococcus
Aminoglycosides
Amphenicols
Cephalosporins
Glycopeptides
Other
Penicillins
Quinolones
Tetracyclines
Escherichia
Aminoglycosides
Amphenicols
Carbapenems
Cephalosporins
Monobactams
Nitrofurans
Other
Penicillins
Quinolones
ulfonamides and Trimethopri
Tetracyclines
Resistance Probability
Age
0.00
0.25
0.50
0.75
1.00
0
25
50
75
100

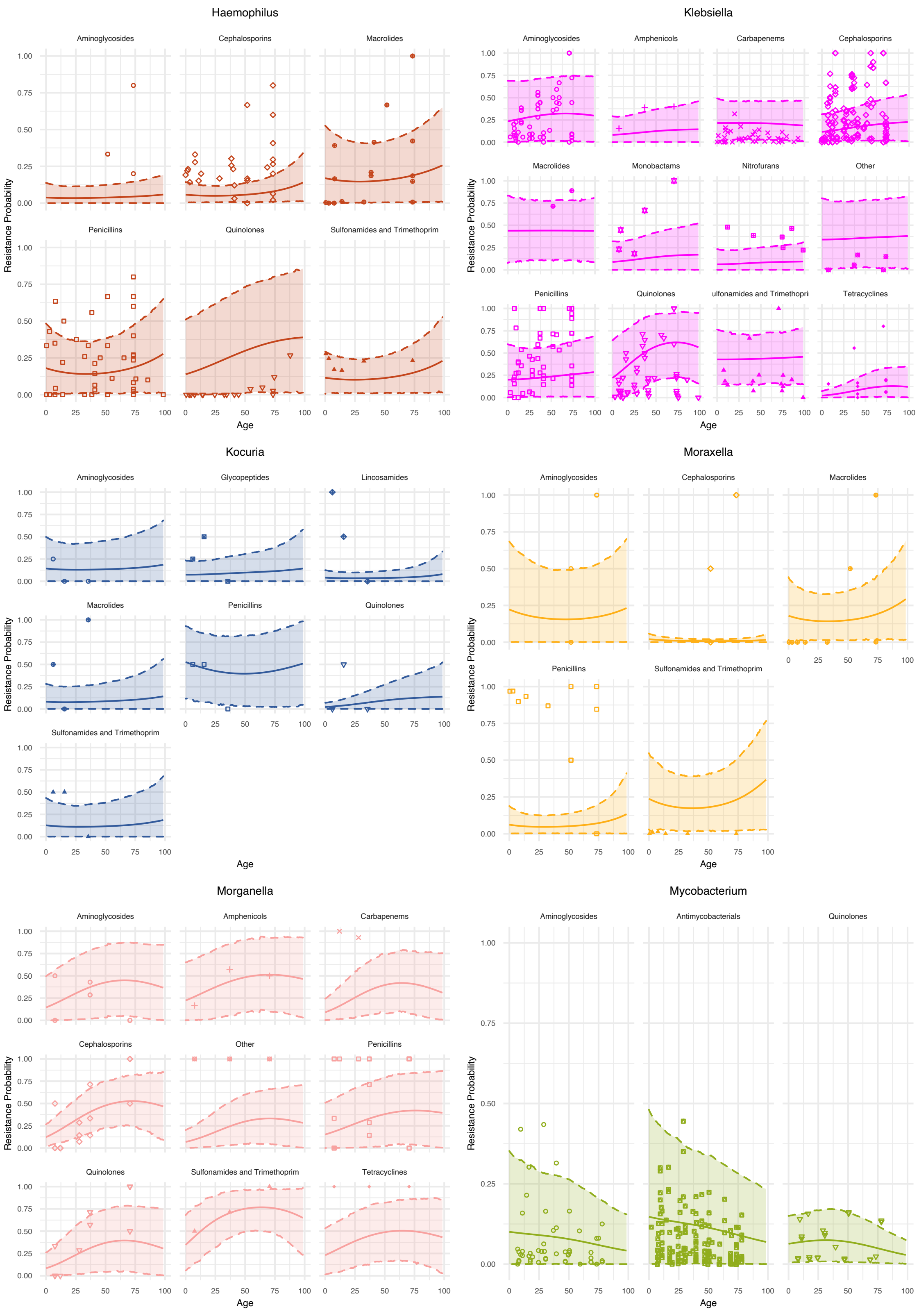
Haemophilus
Aminoglycosides
Cephalosporins
Macrolides
Penicillins
Quinolones
Sulfonamides and Trimethoprim
Klebsiella
Aminoglycosides
Amphenicols
Carbapenems
Cephalosporins
Macrolides
Monobactams
Nitrofurans
Other
Penicillins
Quinolones
ulfonamides and Trimethopri
Tetracyclines
Kocuria
Aminoglycosides
Glycopeptides
Lincosamides
Macrolides
Penicillins
Quinolones
Sulfonamides and Trimethoprim
Moraxella
Aminoglycosides
Cephalosporins
Macrolides
Penicillins
Sulfonamides and Trimethoprim
Morganella
Aminoglycosides
Amphenicols
Carbapenems
Cephalosporins
Other
Penicillins
Quinolones
Sulfonamides and Trimethoprim
Tetracyclines
Mycobacterium
Aminoglycosides
Antimycobacterials
Quinolones
Resistance Probability
Age

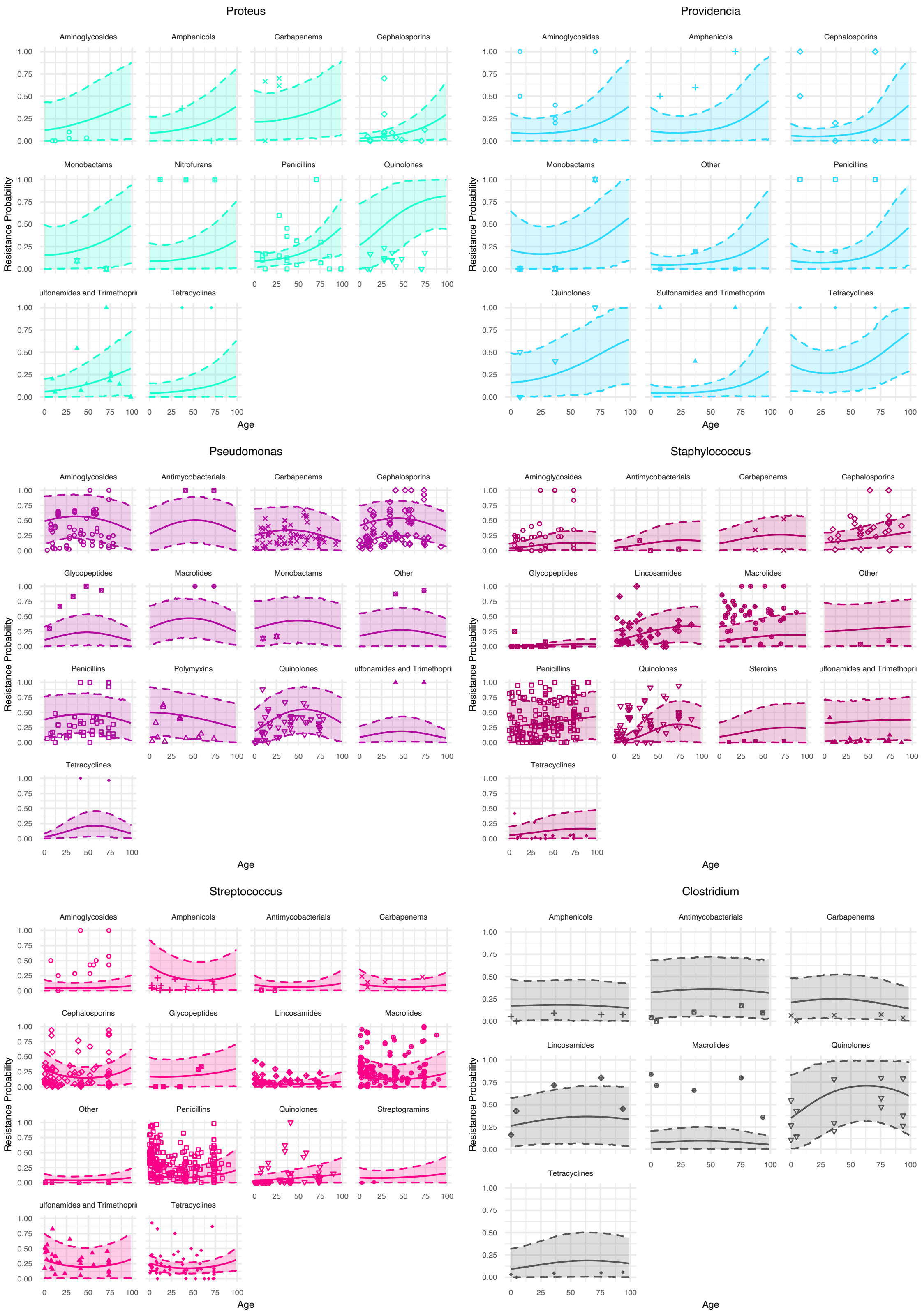
Proteus
Aminoglycosides
Amphenicols
Carbapenems
Cephalosporins
Monobactams
Nitrofurans
Penicillins
Quinolones
ulfonamides and Trimethopri
Tetracyclines
Resistance Probability
Age
Providencia
Aminoglycosides
Amphenicols
Cephalosporins
Monobactams
Other
Penicillins
Quinolones
Sulfonamides and Trimethoprim
Tetracyclines
Resistance Probability
Age
Pseudomonas
Aminoglycosides
Antimycobacterials
Carbapenems
Cephalosporins
Glycopeptides
Macrolides
Monobactams
Other
Penicillins
Polymyxins
Quinolones
ulfonamides and Trimethopri
Tetracyclines
Resistance Probability
Age
Staphylococcus
Aminoglycosides
Antimycobacterials
Carbapenems
Cephalosporins
Glycopeptides
Lincosamides
Macrolides
Other
Penicillins
Quinolones
Steroins
ulfonamides and Trimethopri
Tetracyclines
Resistance Probability
Age
Streptococcus
Aminoglycosides
Amphenicols
Antimycobacterials
Carbapenems
Cephalosporins
Glycopeptides
Lincosamides
Macrolides
Other
Penicillins
Quinolones
Streptogramins
ulfonamides and Trimethopri
Tetracyclines
Resistance Probability
Age
Clostridium
Amphenicols
Antimycobacterials
Carbapenems
Lincosamides
Macrolides
Quinolones
Tetracyclines
Resistance Probability
Age

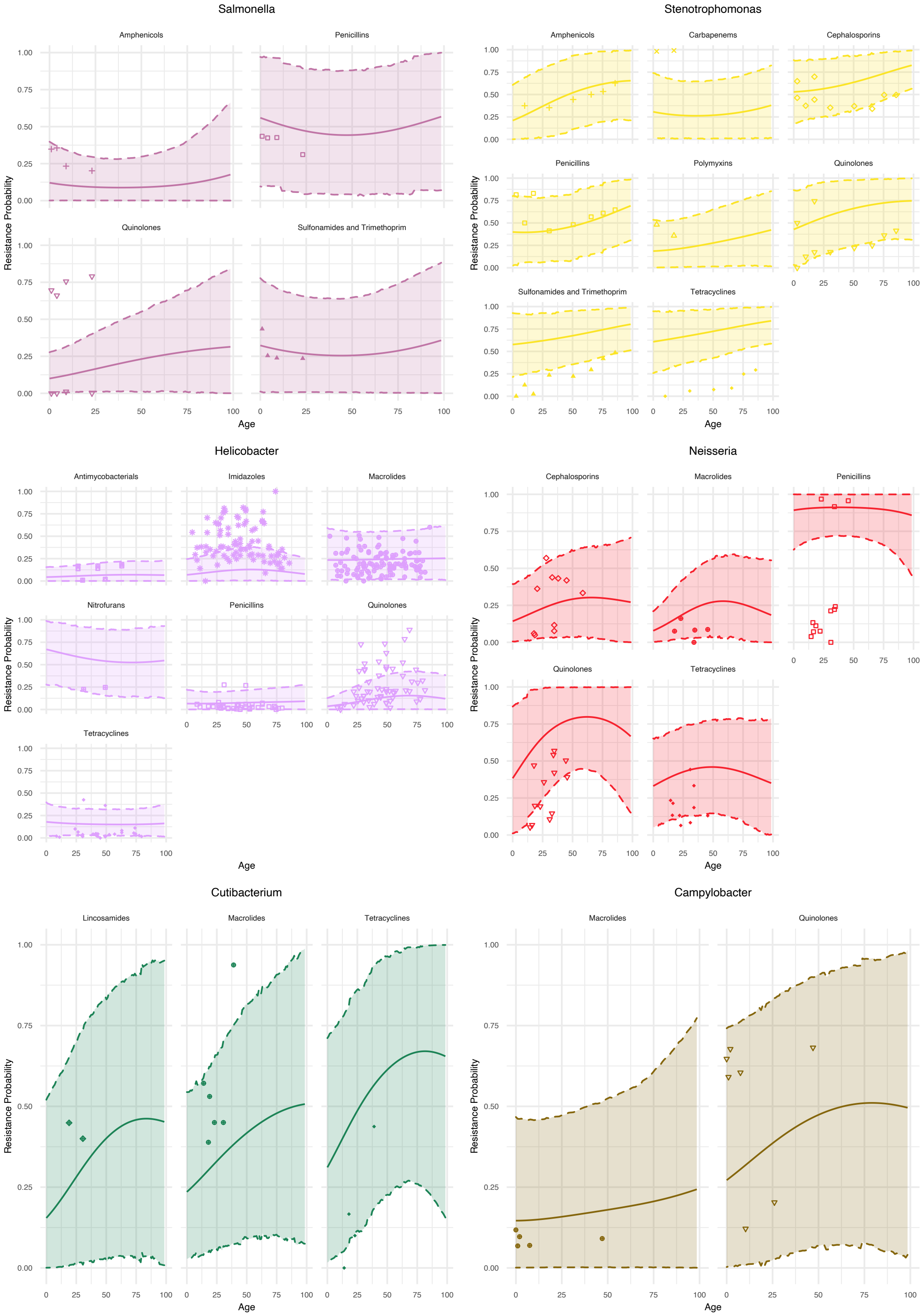
Salmonella
Amphenicols
Penicillins
Quinolones
Sulfonamides and Trimethoprim
Stenotrophomonas
Amphenicols
Carbapenems
Cephalosporins
Penicillins
Polymyxins
Quinolones
Sulfonamides and Trimethoprim
Tetracyclines
Helicobacter
Antimycobacterials
Imidazoles
Macrolides
Nitrofurans
Penicillins
Quinolones
Tetracyclines
Neisseria
Cephalosporins
Macrolides
Penicillins
Quinolones
Tetracyclines
Cutibacterium
Lincosamides
Macrolides
Tetracyclines
Campylobacter
Macrolides
Quinolones
Resistance Probability
Age
0.00
0.25
0.50
0.75
1.00
0
25
50
75
100

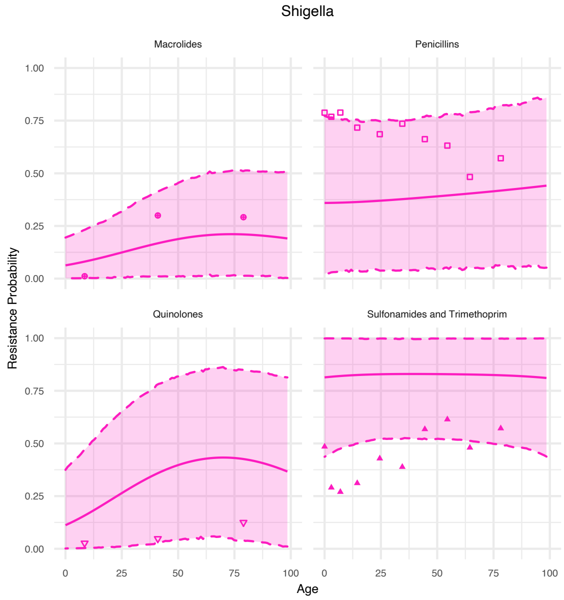


**Fig. S3.** The predicted resistance probability by age with 95% credible intervals for specific antibiotic class and bacterial genera combinations.

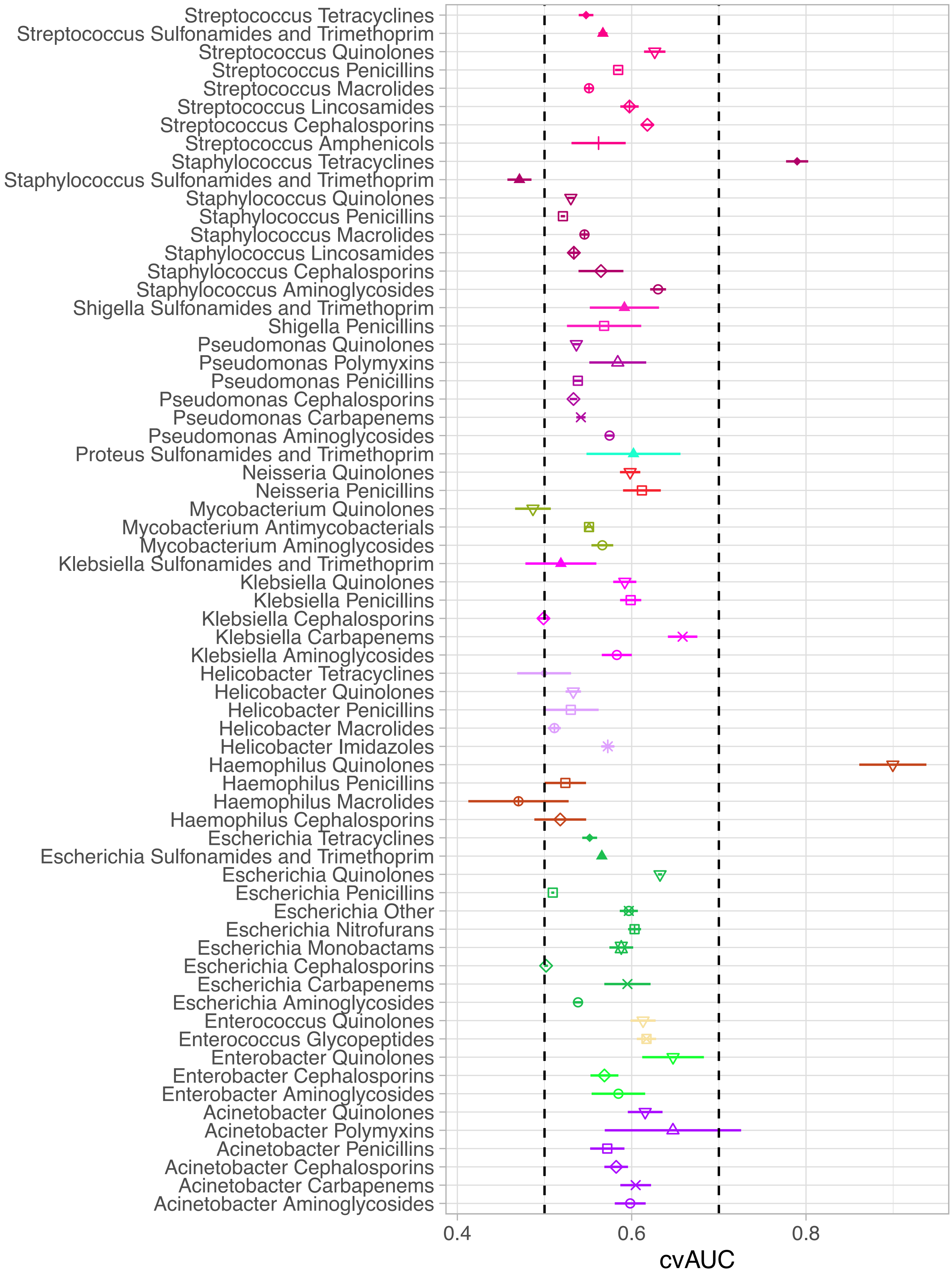
Streptococcus Tetracyclines
Streptococcus Sulfonamides and Trimethoprim
Streptococcus Quinolones
Streptococcus Penicillins
Streptococcus Macrolides
Streptococcus Lincosamides
Streptococcus Cephalosporins
Streptococcus Amphenicols
Staphylococcus Tetracyclines
Staphylococcus Sulfonamides and Trimethoprim
Staphylococcus Quinolones
Staphylococcus Penicillins
Staphylococcus Macrolides
Staphylococcus Lincosamides
Staphylococcus Cephalosporins
Staphylococcus Aminoglycosides
Shigella Sulfonamides and Trimethoprim
Shigella Penicillins
Pseudomonas Quinolones
Pseudomonas Polymyxins
Pseudomonas Penicillins
Pseudomonas Cephalosporins
Pseudomonas Carbapenems
Pseudomonas Aminoglycosides
Proteus Sulfonamides and Trimethoprim
Neisseria Quinolones
Neisseria Penicillins
Mycobacterium Quinolones
Mycobacterium Antimycobacterials
Mycobacterium Aminoglycosides
Klebsiella Sulfonamides and Trimethoprim
Klebsiella Quinolones
Klebsiella Penicillins
Klebsiella Cephalosporins
Klebsiella Carbapenems
Klebsiella Aminoglycosides
Helicobacter Tetracyclines
Helicobacter Quinolones
Helicobacter Penicillins
Helicobacter Macrolides
Helicobacter Imidazoles
Haemophilus Quinolones
Haemophilus Penicillins
Haemophilus Macrolides
Haemophilus Cephalosporins
Escherichia Tetracyclines
Escherichia Sulfonamides and Trimethoprim
Escherichia Quinolones
Escherichia Penicillins
Escherichia Other
Escherichia Nitrofurans
Escherichia Monobactams
Escherichia Cephalosporins
Escherichia Carbapenems
Escherichia Aminoglycosides
Enterococcus Quinolones
Enterococcus Glycopeptides
Enterobacter Quinolones
Enterobacter Cephalosporins
Enterobacter Aminoglycosides
Acinetobacter Quinolones
Acinetobacter Polymyxins
Acinetobacter Penicillins
Acinetobacter Cephalosporins
Acinetobacter Carbapenems
Acinetobacter Aminoglycosides
0.4
0.6
0.8
cvAUC

**Fig. S4.** Plot showing the 10-fold cross-validated Area Under the ROC Curve (cvAUC) for antibiotic-bacteria combinations with resistance probability for 10 or more age categories. Each point represents the cvAUC for each combination, with lines representing 95% confidence intervals. Vertical dashed lines show cvAUC = 0.5 (random) and cvAUC = 0.7 ("good" prediction).

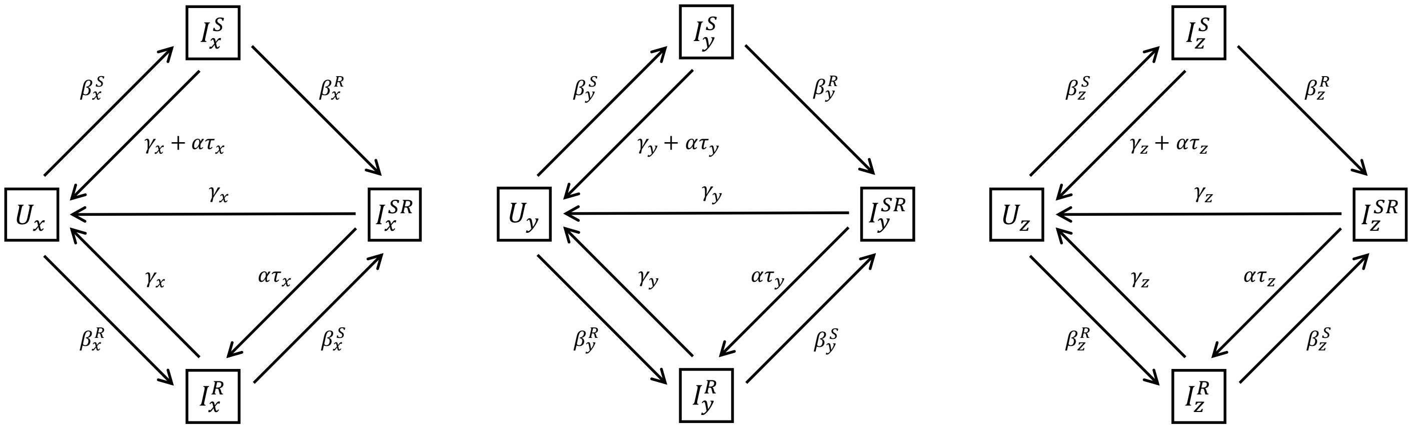


**Fig. S5.** Diagram of the susceptible-infected compartmental model used to describe the dynamics of resistance for three distinct but interacting sub-populations (Children, x; Adults, y; and Elderly, z).

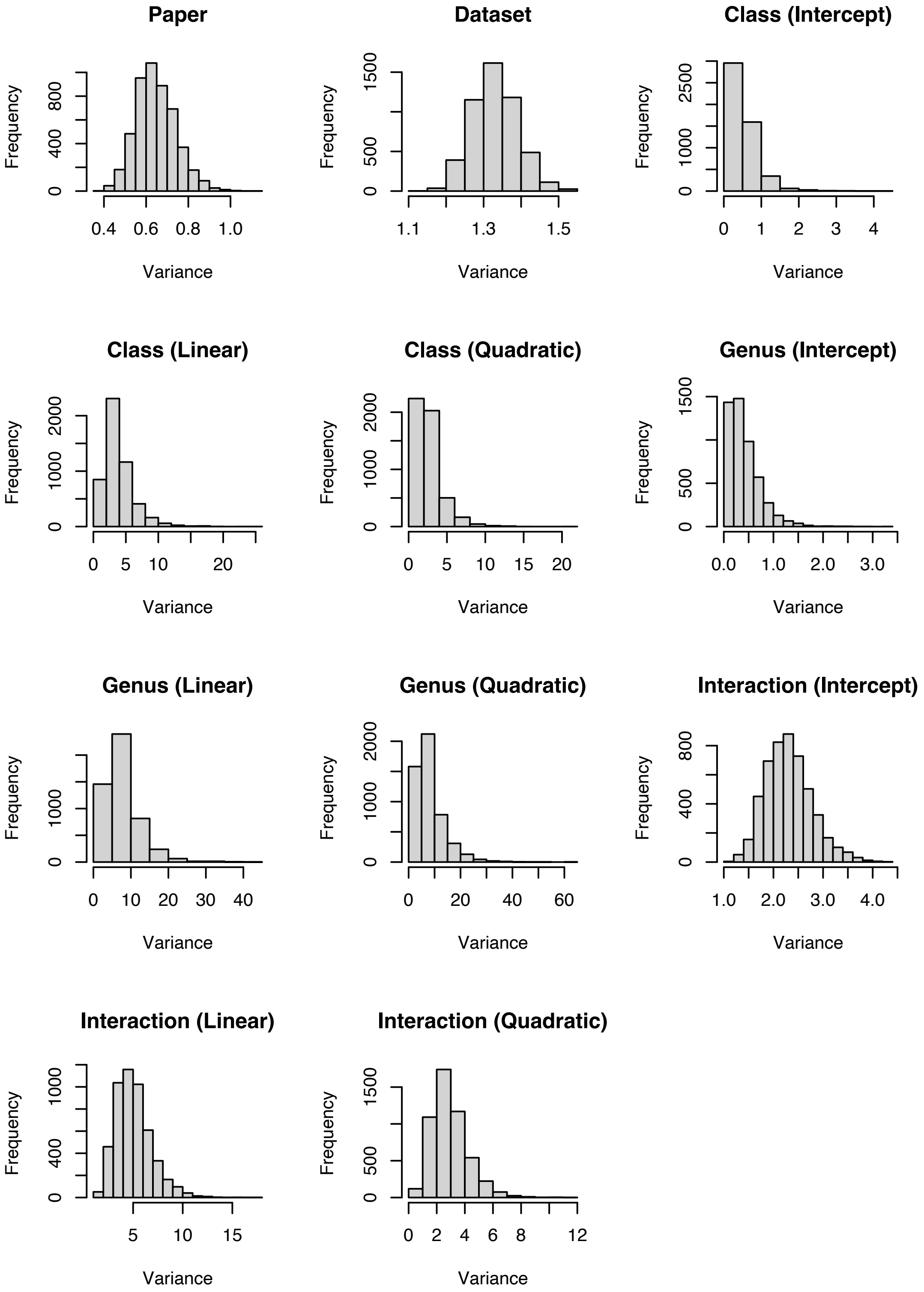


**Fig. S6.** Histogram of the posterior distribution of each random effect to assess the significance of random effects.

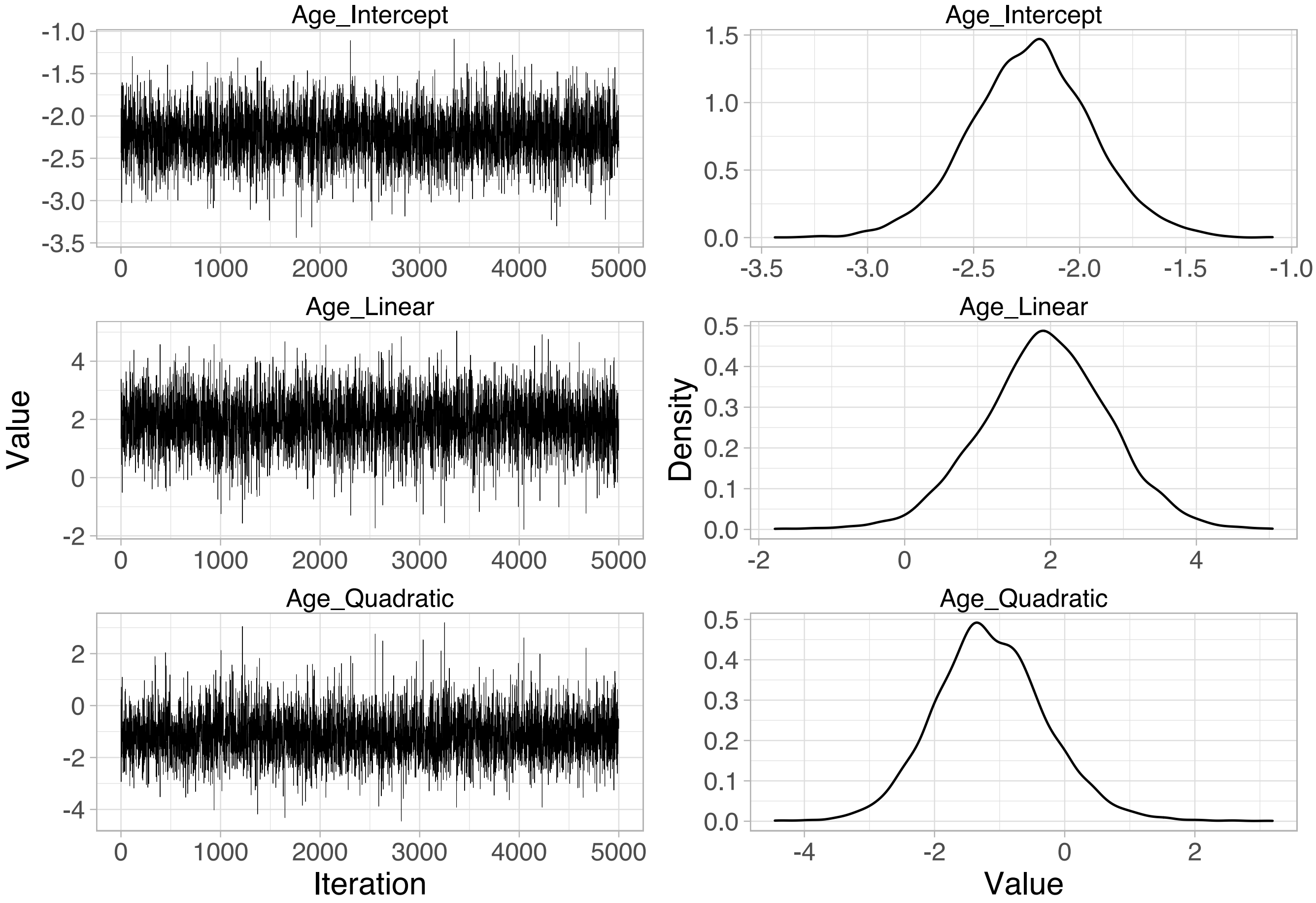


**Fig. S7.** Trace and density estimate for the fixed effects (top = Intercept, middle = Age, bottom = Age$^2$) to assess model convergence.

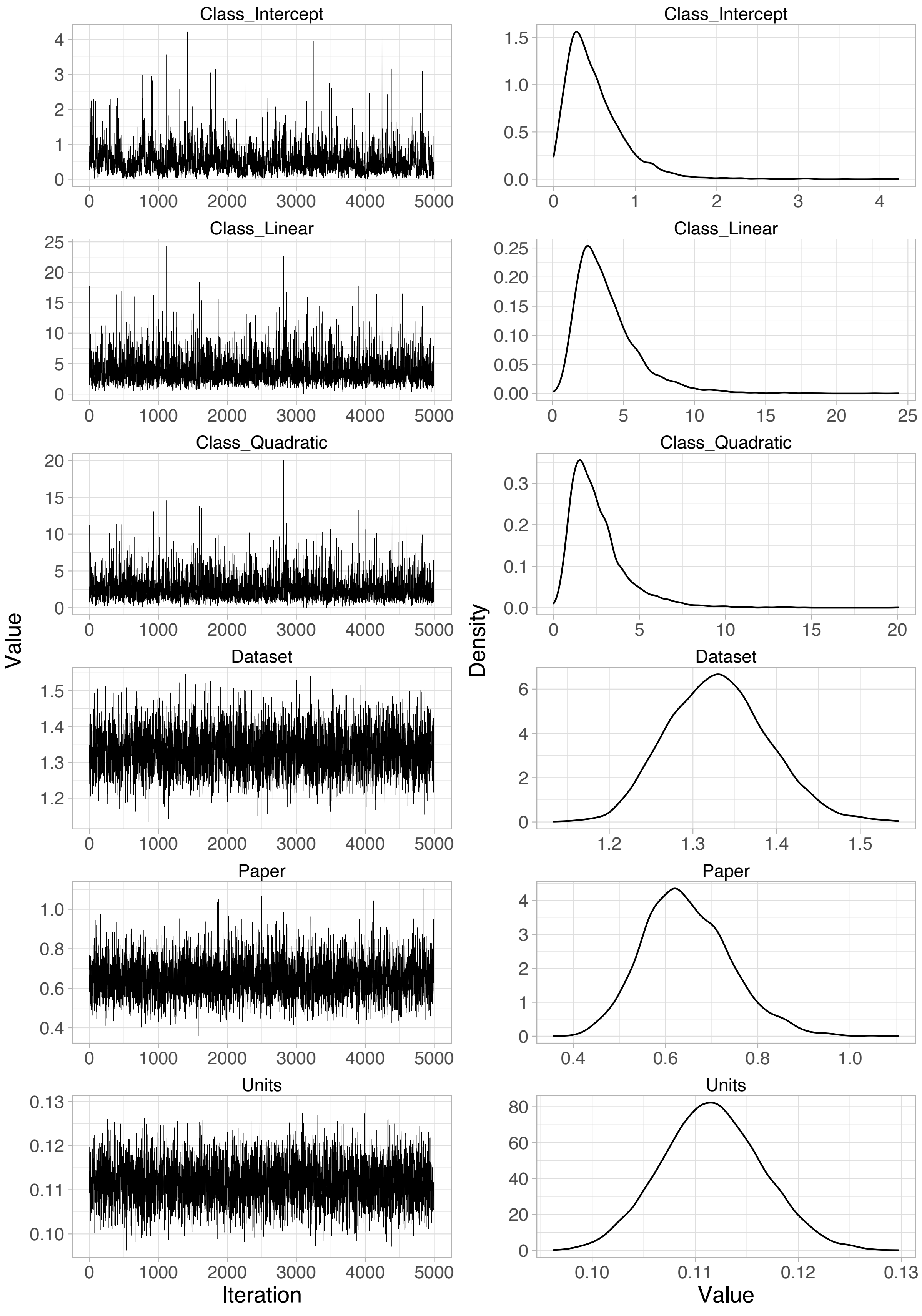
Class_Intercept
Class_Intercept
Class_Linear
Class_Linear
Class_Quadratic
Class_Quadratic
Dataset
Dataset
Paper
Paper
Units
Units
Value
Density
Iteration
Value

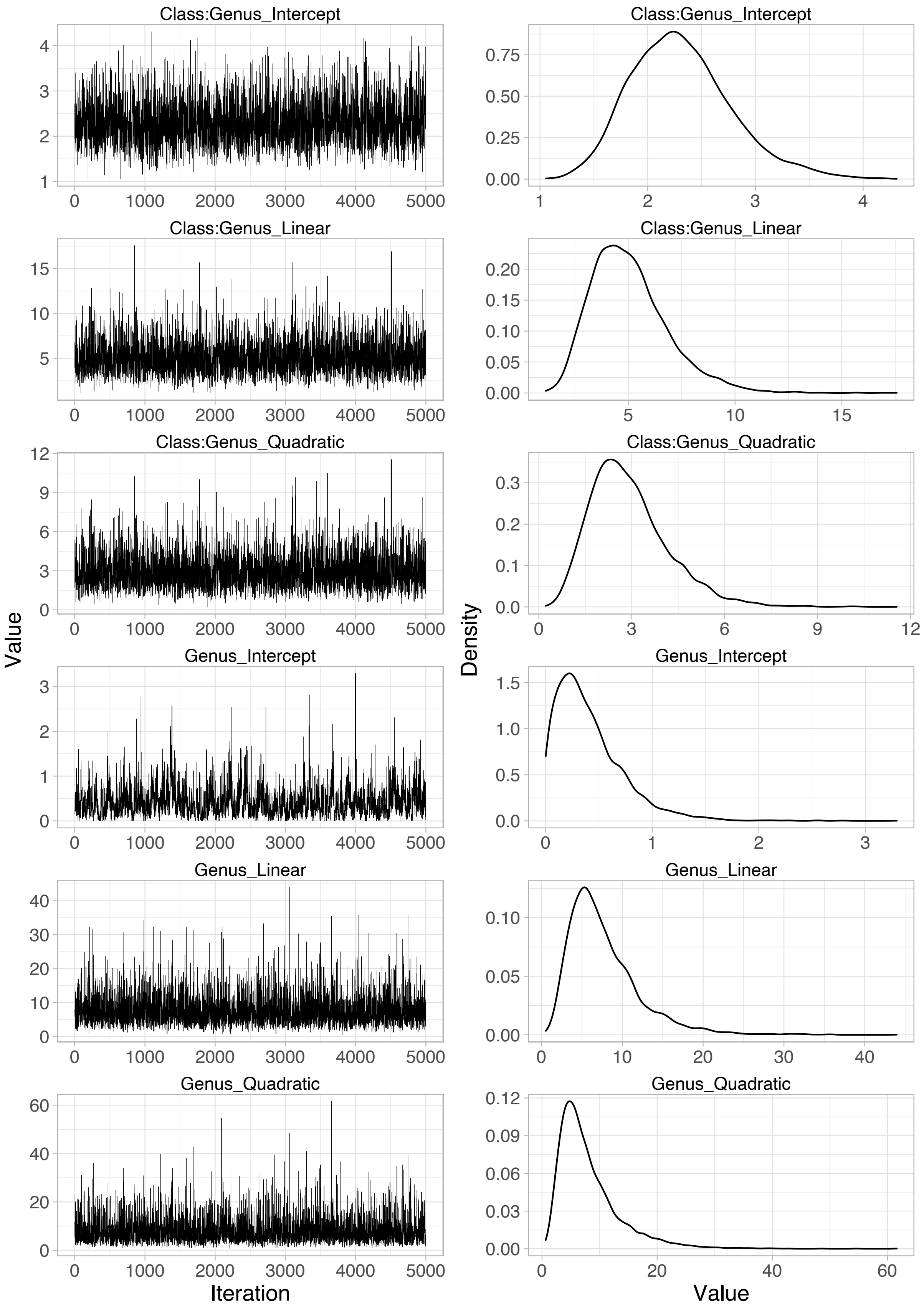

Class:Genus_Intercept
Class:Genus_Intercept
Class:Genus_Linear
Class:Genus_Linear
Class:Genus_Quadratic
Class:Genus_Quadratic
Genus_Intercept
Genus_Intercept
Genus_Linear
Genus_Linear
Genus_Quadratic
Genus_Quadratic
Value
Iteration
Density
Value

**Fig. S8.** Trace and density estimate for the random effects (Paper (Intercept), Dataset (Intercept), Class (Intercept), Class (Linear), Class (Quadratic), Genus (Intercept), Genus (Linear), Genus (Quadratic), Class:Genus (Intercept), Class:Genus (Linear), Class:Genus (Quadratic)) to assess model convergence.

| PMID | | | | | | | |
|---|---|---|---|---|---|---|---|
| 32576154 | 21412585 | 11414411 | 24618888 | 8821601 | 23840002 | 20211888 | 20814127 |
| 31798838 | 21131751 | 11091043 | 24094837 | 7623880 | 23433241 | 21294447 | 11095556 |
| 31499177 | 20638598 | 10952606 | 23746128 | 7795081 | 22572670 | 20966623 | 9239768 |
| 30845149 | 20652344 | 10797084 | 23786942 | 3885842 | 21873378 | 18808361 | 26701944 |
| 30819135 | 20225774 | 10629012 | 23540793 | 30947695 | 20092664 | 17083381 | 17061600 |
| 29961840 | 19751432 | 10413735 | 21450969 | 29982570 | 18356156 | 16174685 | 27244961 |
| 29284419 | 19297267 | 9195057 | 19074658 | 30550238 | 18313513 | 11777360 | 20487614 |
| 28961888 | 18397923 | 8592999 | 19030711 | 26740324 | 15947850 | 11713400 | 16351027 |
| 29138101 | 18227764 | 7619999 | 18077618 | 25013979 | 15609696 | 10619736 | 18197977 |
| 28949279 | 17977993 | 7893874 | 18073151 | 24631369 | 11597287 | 31188848 | 24758533 |
| 28615465 | 17893757 | 2052962 | 17974221 | 23573984 | 9498447 | 30440065 | 16506084 |
| 28506722 | 17105833 | 2496174 | 17933699 | 21901873 | 9301992 | 29992016 | 16212123 |
| 27744103 | 16766005 | 3655402 | 17539982 | 21545312 | 8824968 | 28501582 | 11173812 |
| 27631162 | 16549015 | 6341595 | 17530038 | 19808237 | 31828045 | 26833774 | 7963641 |
| 26918818 | 16473503 | 12851 | 16520472 | 19703843 | 31547889 | 26577970 | 20038489 |
| 26937912 | 15715717 | 31582419 | 16023881 | 19269873 | 31440915 | 26842176 | 27048586 |
| 26667678 | 15573843 | 31529852 | 15504260 | 17728510 | 31385490 | 23959315 | 21220537 |
| 26147515 | 15214879 | 30917150 | 15328089 | 17335597 | 30805183 | 19446980 | 11516934 |
| 25525198 | 14986245 | 30203738 | 15265833 | 16102308 | 31357057 | 19287838 | 11343444 |
| 25220697 | 14614074 | 30646267 | 15215091 | 15310167 | 29597292 | 14553870 | 30976388 |
| 24652673 | 14585850 | 30041525 | 12925100 | 15075018 | 28480653 | 12018473 | 15128722 |
| 23725981 | 12458134 | 28681811 | 12792711 | 12760850 | 26987479 | 10619737 | 20013094 |
| 24156217 | 12234869 | 28098543 | 12684907 | 12667237 | 25781009 | 10543737 | 17888610 |
| 23587776 | 12096005 | 27896620 | 12542200 | 12363009 | 25759550 | 16529904 | 16598044 |
| 23090037 | 11959559 | 27306116 | 11858909 | 11136262 | 25814302 | 11231185 | 17577813 |
| 22795635 | 11936736 | 27178769 | 11731940 | 30005979 | 25504299 | 6202743 | |
| 22579652 | 11932142 | 26980934 | 11679555 | 29969753 | 25581808 | 30010391 | |
| 22217142 | 11389500 | 26747068 | 10868529 | 29468052 | 25413920 | 26243258 | |
| 22103749 | 11353617 | 26632819 | 9847953 | 28839298 | 20943819 | 26302942 | |
| 21880098 | 11158087 | 25661133 | 9182104 | 24687499 | 20809835 | 23625165 | |

**Table S1.** List of PMIDs of studies included in the meta-analysis.

| | Reason for exclusion |
|---|---|
| Abstract Screen | Antibiotic prescribing practises, stewardship, or consumption |
| | Age as a risk factor for frequency of infection (regardless of resistance status) |
| | Resistant infection as a risk factor for outcome (e.g., death, morbidity, treatment failure, infection following colonisation) |
| | Only include other risk factors for probability of resistant infection (e.g., gender, location) |
| | Risk factors for sub clones associated with resistance (e.g., Beijing genotype of Mycobacterium tuberculosis) |
| | Risk factors for another variable in resistant infections (e.g., risk factors for bacteraemia in MDR-AB, risk factors for infection versus colonisation with CRE) |
| | Prevalence of specific resistant infection (e.g., MRSA, MDR-AB, CRE, VRE, ESBL) |
| | Outbreak or transmission dynamics of a specific drug resistant clone |
| | Risk factors for specific type of resistance within resistant population |
| Full Text Screen | No full text access |
| | No extractable data<br>- Statistical difference between the average host age of resistant and sensitive infections<br>- Qualitative statements only<br>- Odds ratios only<br>- Sample size per age category not reported (% only)<br>- Comparison of average MIC between age categories<br>- Data inconsistencies (e.g., obvious mistakes)<br>- Extrapolated estimates for resistance probability<br>- Age categories not properly defined (e.g., “Adult”, “Child”) |
| | Invalid comparison (host age as a risk factor for probability of…)<br>- Resistant infection compared to no infection (e.g., MRSA vs no MRSA)<br>- Resistant infection compared to non-resistant infection caused by a different pathogen (e.g., MDR-AB vs non-MDR GNO)<br>- Mono-resistant infection compared to non-mono-resistant infection (e.g., Isoniazid monoresistance vs other)<br>- Resistant infection compared to pan-sensitive infection<br>- Age categories not comparable (e.g., symptomatic adults vs asymptomatic children) |
| | Non-random sampling<br>- Only include resistant infections (e.g., other resistance in MRSA)<br>- Case-control study with patients selected based on resistance status of infection (e.g., all resistant infections + next admitted sensitive infection) |
| | Resistance determined by genotype |
| | Acquired resistance (i.e., where the same infection was sensitive at the first test and resistant at the second test) |
| | Outbreaks (e.g., MRSA) |
| | Non-specific bacterial species or bacterial genus, non-specific antibiotic or antibiotic class, or multi-drug resistance |

**Table S2.** Reasons for exclusion of articles based on abstract and full text screening.

| Bacterial Genus | Bacteria |
|---|---|
| Acinetobacter | *baumannii* |
| Aerococcus | |
| Campylobacter | |
| Citrobacter | *freundii* |
| Clostridium | *dificile* |
| Cutibacterium | *acnes* |
| Enterobacter | *cloacae* |
| Enterococcus | *faecalis; faecium* |
| Escherichia | *coli* |
| Haemophilus | *influenzae* |
| Helicobacter | *pylori* |
| Klebsiella | *oxytoca; pneumoniae* |
| Kocuria | |
| Moraxella | *catarrhalis* |
| Morganella | *morganii* |
| Mycobacterium | *tuberculosis* |
| Neisseria | *gonorrhoeae* |
| Proteus | *mirabilis; vulgaris* |
| Providencia | |
| Pseudomonas | *aeruginosa* |
| Salmonella | |
| Shigella | |
| Staphylococcus | CoNS*; aureus; epidermidis; saprophyticus* |
| Stenotrophomonas | *maltophila* |
| Streptococcus | *agalactiae; pneumoniae; pyogenes* |

**Table S3.** All bacteria included in the meta-analysis and their grouping by genus.

| Antibiotic Class | Antibiotic |
|---|---|
| Aminoglycosides (J01G) | amikacin; gentamicin; kanamycin; netilmicin; streptomycin; tobramycin |
| Amphenicols (J01B) | chloramphenicol |
| Anitmycobacterials (J04A) | 4-aminosalicylic acid; capreomycin; cycloserine; ethambutol; ethionamide; isoniazid; protionamide; pyrazinamide; rifabutin; rifampicin |
| Carbapenems (J01DH) | doripenem; ertapenem; imipenem and cilastatin; meropenem |
| Cephalosporins (J01DB, J01DC, J01DD, J01DE) | cefaclor; cefalexin; cefalotin; cefazolin; cefepime; cefixime; cefoperazone; cefoperazone and beta-lactamase inhibitor; cefotaxime; cefoxitin; cefpodoxime; cefprozil; ceftazidime; ceftriaxone; cefuroxime |
| Glycopeptides (J01XA) | teicoplanin; vancomycin |
| Imidazoles (J01XD) | metronidazole |
| Lincosamides (J01FF) | clindamycin |
| Macrolides (J01FA) | azithromycin; clarithromycin; erythromycin; telithromycin |
| Monobactams (J01DF) | aztreonam |
| Nitrofurans (J01XE) | furazolidone; nitrofurantoin |
| Other (J01XX) | fosfomycin; linezolid |
| Penicillins (J01C) | amoxicillin; amoxicillin and beta-lactamase inhibitor; ampicillin; ampicillin and beta-lactamase inhibitor; benzylpenicillin; carbenicillin; methicillin; mezlocillin; oxacillin; phenoxymethylpenicillin; piperacillin; piperacillin and beta-lactamase inhibitor; ticarcillin; ticarcillin and beta-lactamase inhibitor |
| Polymyxins (J01XB) | colistin; polymyxin B |
| Quinolones (J01M) | ciprofloxacin; enoxacin; garenoxacin; gatifloxacin; levofloxacin; moxifloxacin; nalidixic acid; norfloxacin; ofloxacin; sparfloxacin |
| Steroids (J01XC) | fusidic acid |
| Streptogramins (J01FG) | quinupristin/dalfopristin |
| Sulfonamides and trimethoprim (J01E) | sulfamethoxazole; sulfamethoxazole and trimethoprim; sulfonamide; trimethoprim |
| Tetracyclines (J01A) | minocycline; sulfonamide; tetracycline; tigecycline |

**Table S4.** All antibiotics included in the meta-analysis and their grouping by antibiotic class.

| | Symbol | Definition | Value | Range | Notes |
|---|---|---|---|---|---|
| **Variables** | $U$ | Uninfected | | | |
| | $I^S$ | Infected with antibiotic-sensitive bacteria | | | |
| | $I^R$ | Infected with antibiotic-resistant bacteria | | | |
| | $I^{SR}$ | Co-infected | | | |
| | $x$ | Child sub-population | | | |
| | $y$ | Adult sub-population | | | |
| | $z$ | Elderly sub-population | | | |
| **Parameters** | $\beta$ | Transmission rate | 1/24 $\text{day}^{-1}$ | 1/27 – 1/21 | Transmission rate was calculated by $\frac{\gamma}{(1-\text{prevalence})}$ with prevalence (across all ages) estimated as 0.2 [34,50–53] |
| | $ct$ | Transmission cost of resistance | 0.05 | | Cost of resistance was estimated [54–56] |
| | $\gamma$ | Immune clearance rate | 1/30 $\text{day}^{-1}$ | 1/34 – 1/26 | Immune clearance rate was calculated by $\frac{1}{\text{carriage duration}}$ with untreated carriage duration (across all ages) estimated as 30 days [48,49] |
| | $\alpha$ | Antibiotic clearance rate | 1 $\text{day}^{-1}$ | | Antibiotic clearance was calculated by $\frac{1}{\text{infection duration}}$ with treated infection duration estimated as 1 day [57] |
| | $\tau$ | Antibiotic treatment rate | 0.001 $\text{day}^{-1}$ | 0 – 0.002 | Treatment rate was estimated using data from the European Centre for Disease Prevention and Control (ECDC) on the primary-care consumption of penicillins in the UK in 2019 [58]. 11.12 Defined Daily Doses (DDDs) per 1000 persons per day was converted to treatment rate by assuming that 10 DDDs comprise 1 treatment course [59]. |
| | $m$ | Between sub-population mixing | | 0.01, 0.05 | |

**Table S5.** Definition and value of each parameter used in the susceptible-infected compartmental model. Range gives the upper and lower bounds when that parameter was varied.

| Age | Mixing | Treatment | Transmission | Clearance | Prevalence | Resistance |
|---|---|---|---|---|---|---|
| Child | 0.01 | 0 | 0.4166667 | 0.3333333 | 0.19564594 | 0.04683356 |
| Adult | 0.01 | 0.1 | 0.4166667 | 0.3333333 | 0.17500624 | 0.08507261 |
| Elderly | 0.01 | 0.2 | 0.4166667 | 0.3333333 | 0.1572718 | 0.17673058 |
| Child | 0.01 | 0.2 | 0.4166667 | 0.3333333 | 0.1572718 | 0.17673058 |
| Adult | 0.01 | 0.1 | 0.4166667 | 0.3333333 | 0.17500624 | 0.08507261 |
| Elderly | 0.01 | 0 | 0.4166667 | 0.3333333 | 0.19564594 | 0.04683356 |
| Child | 0.01 | 0.05 | 0.4166667 | 0.3333333 | 0.18518106 | 0.05872211 |
| Adult | 0.01 | 0.2 | 0.4166667 | 0.3333333 | 0.15720451 | 0.17099124 |
| Elderly | 0.01 | 0.05 | 0.4166667 | 0.3333333 | 0.18518106 | 0.05872211 |
| Child | 0.01 | 0.125 | 0.4166667 | 0.3333333 | 0.17023008 | 0.0795721 |
| Adult | 0.01 | 0.05 | 0.4166667 | 0.3333333 | 0.18540563 | 0.04636777 |
| Elderly | 0.01 | 0.125 | 0.4166667 | 0.3333333 | 0.17023008 | 0.0795721 |
| Child | 0.01 | 0.1 | 0.3703704 | 0.3333333 | 0.09479146 | 0.14250338 |
| Adult | 0.01 | 0.1 | 0.4166667 | 0.3333333 | 0.17469905 | 0.1467118 |
| Elderly | 0.01 | 0.1 | 0.4761905 | 0.3333333 | 0.26938477 | 0.2299572 |
| Child | 0.01 | 0.1 | 0.4761905 | 0.3333333 | 0.26938477 | 0.2299572 |
| Adult | 0.01 | 0.1 | 0.4166667 | 0.3333333 | 0.17469905 | 0.1467118 |
| Elderly | 0.01 | 0.1 | 0.3703704 | 0.3333333 | 0.09479146 | 0.14250338 |
| Child | 0.01 | 0.1 | 0.3921569 | 0.3333333 | 0.13150307 | 0.1337457 |
| Adult | 0.01 | 0.1 | 0.4761905 | 0.3333333 | 0.26925407 | 0.22638361 |
| Elderly | 0.01 | 0.1 | 0.3921569 | 0.3333333 | 0.13150307 | 0.1337457 |
| Child | 0.01 | 0.1 | 0.4444444 | 0.3333333 | 0.22148175 | 0.13884966 |
| Adult | 0.01 | 0.1 | 0.3703704 | 0.3333333 | 0.09521692 | 0.10120842 |
| Elderly | 0.01 | 0.1 | 0.4444444 | 0.3333333 | 0.22148175 | 0.13884966 |
| Child | 0.01 | 0.1 | 0.4166667 | 0.375 | 0.09630101 | 0.14753436 |
| Adult | 0.01 | 0.1 | 0.4166667 | 0.3333333 | 0.17454798 | 0.15562666 |
| Elderly | 0.01 | 0.1 | 0.4166667 | 0.2916667 | 0.2670822 | 0.2532204 |
| Child | 0.01 | 0.1 | 0.4166667 | 0.2916667 | 0.2670822 | 0.2532204 |
| Adult | 0.01 | 0.1 | 0.4166667 | 0.3333333 | 0.17454798 | 0.15562666 |
| Elderly | 0.01 | 0.1 | 0.4166667 | 0.375 | 0.09630101 | 0.14753436 |
| Child | 0.01 | 0.1 | 0.4166667 | 0.3541667 | 0.1324115 | 0.13899909 |
| Adult | 0.01 | 0.1 | 0.4166667 | 0.2916667 | 0.26695699 | 0.24902941 |
| Elderly | 0.01 | 0.1 | 0.4166667 | 0.3541667 | 0.1324115 | 0.13899909 |
| Child | 0.01 | 0.1 | 0.4166667 | 0.3125 | 0.22015331 | 0.15199478 |
| Adult | 0.01 | 0.1 | 0.4166667 | 0.375 | 0.09683006 | 0.10482265 |
| Elderly | 0.01 | 0.1 | 0.4166667 | 0.3125 | 0.22015331 | 0.15199478 |
| Child | 0.05 | 0 | 0.4166667 | 0.3333333 | 0.18928791 | 0.05112385 |
| Adult | 0.05 | 0.1 | 0.4166667 | 0.3333333 | 0.17549844 | 0.06101896 |
| Elderly | 0.05 | 0.2 | 0.4166667 | 0.3333333 | 0.16300146 | 0.07247446 |

| | | | | | | |
|---|---|---|---|---|---|---|
| Child | 0.05 | 0.2 | 0.4166667 | 0.3333333 | 0.16300146 | 0.07247446 |
| Adult | 0.05 | 0.1 | 0.4166667 | 0.3333333 | 0.17549844 | 0.06101896 |
| Elderly | 0.05 | 0 | 0.4166667 | 0.3333333 | 0.18928791 | 0.05112385 |
| Child | 0.05 | 0.05 | 0.4166667 | 0.3333333 | 0.18215539 | 0.05591909 |
| Adult | 0.05 | 0.2 | 0.4166667 | 0.3333333 | 0.1629247 | 0.07253274 |
| Elderly | 0.05 | 0.05 | 0.4166667 | 0.3333333 | 0.18215539 | 0.05591909 |
| Child | 0.05 | 0.125 | 0.4166667 | 0.3333333 | 0.17200224 | 0.06402414 |
| Adult | 0.05 | 0.05 | 0.4166667 | 0.3333333 | 0.18198076 | 0.05612523 |
| Elderly | 0.05 | 0.125 | 0.4166667 | 0.3333333 | 0.17200224 | 0.06402414 |
| Child | 0.05 | 0.1 | 0.3703704 | 0.3333333 | 0.12858673 | 0.12931183 |
| Adult | 0.05 | 0.1 | 0.4166667 | 0.3333333 | 0.17878198 | 0.13225496 |
| Elderly | 0.05 | 0.1 | 0.4761905 | 0.3333333 | 0.24883522 | 0.14619245 |
| Child | 0.05 | 0.1 | 0.4761905 | 0.3333333 | 0.24883522 | 0.14619245 |
| Adult | 0.05 | 0.1 | 0.4166667 | 0.3333333 | 0.17878198 | 0.13225496 |
| Elderly | 0.05 | 0.1 | 0.3703704 | 0.3333333 | 0.12858673 | 0.12931183 |
| Child | 0.05 | 0.1 | 0.3921569 | 0.3333333 | 0.15026679 | 0.12319067 |
| Adult | 0.05 | 0.1 | 0.4761905 | 0.3333333 | 0.24799835 | 0.13914419 |
| Elderly | 0.05 | 0.1 | 0.3921569 | 0.3333333 | 0.15026679 | 0.12319067 |
| Child | 0.05 | 0.1 | 0.4444444 | 0.3333333 | 0.21079396 | 0.10385877 |
| Adult | 0.05 | 0.1 | 0.3703704 | 0.3333333 | 0.12790543 | 0.09651313 |
| Elderly | 0.05 | 0.1 | 0.4444444 | 0.3333333 | 0.21079396 | 0.10385877 |
| Child | 0.05 | 0.1 | 0.4166667 | 0.375 | 0.12927025 | 0.13968187 |
| Adult | 0.05 | 0.1 | 0.4166667 | 0.3333333 | 0.17843049 | 0.14422949 |
| Elderly | 0.05 | 0.1 | 0.4166667 | 0.2916667 | 0.24662357 | 0.16211369 |
| Child | 0.05 | 0.1 | 0.4166667 | 0.2916667 | 0.24662357 | 0.16211369 |
| Adult | 0.05 | 0.1 | 0.4166667 | 0.3333333 | 0.17843049 | 0.14422949 |
| Elderly | 0.05 | 0.1 | 0.4166667 | 0.375 | 0.12927025 | 0.13968187 |
| Child | 0.05 | 0.1 | 0.4166667 | 0.3541667 | 0.15060442 | 0.13295356 |
| Adult | 0.05 | 0.1 | 0.4166667 | 0.2916667 | 0.2458138 | 0.15379731 |
| Elderly | 0.05 | 0.1 | 0.4166667 | 0.3541667 | 0.15060442 | 0.13295356 |
| Child | 0.05 | 0.1 | 0.4166667 | 0.3125 | 0.20964667 | 0.11273367 |
| Adult | 0.05 | 0.1 | 0.4166667 | 0.375 | 0.12872141 | 0.10256716 |
| Elderly | 0.05 | 0.1 | 0.4166667 | 0.3125 | 0.20964667 | 0.11273367 |

**Table S6.** Parameter values for each run of the susceptible-infected compartmental model, along with prevalence of infection and proportion of resistant infections.
